# Determinants of DNA-sequence-based Diagnostic Yield in the CSER Consortium

**DOI:** 10.64898/2026.04.20.26351140

**Authors:** Yusuph Mavura, David Crosslin, Kathleen Ferar, James MJ Lawlor, John M. Greally, Lucia Hindorff, Gail P Jarvik, Sara Kalla, Barbara A Koenig, Mark Kvale, Pui-Yan Kwok, Mary Norton, Sharon E. Plon, Bradford C. Powell, Anne Slavotinek, Michelle L Thompson, Alice B Popejoy, Eimear E. Kenny, Neil Risch

**Affiliations:** Department of Biomedical Data Science, Stanford University, Palo Alto, CA; BioMed. Informatics and Genomics, John W. Deming Dept. of Med., Tulane Univ. Sch. of Med., New Orleans, LA; Department of Biomedical Informatics and Medical Education, University of Washington School of Medicine, Seattle, WA; HudsonAlpha Inst. for Biotechnology, Huntsville, AL; Departments of Genetics and Pediatrics, Albert Einstein College of Medicine, Bronx, NY; NHGRI, NIH, Bethesda, MD; Medicine (Medical Genetics) and Genome Sciences, Univ. of Washington Med. Ctr., Seattle, WA; HGSC - Baylor Coll. of Med., Houston, TX; Institute for Human Genetics, University of California San Francisco, San Francisco, CA; Program in Bioethics, University of California San Francisco, San Francisco, CA; Cardiovascular Research Institute and Department of Dermatology, University of California San Francisco, San Francisco, CA; Division of Maternal Fetal Medicine, Department of Obstetrics, Gynecology, and Reproductive Sciences, University of California, San Francisco, San Francisco CA; Molecular and Human Genetics Dept., Baylor Coll. Med., Houston, TX; Pediatrics-Hematology-Oncology Dept, Baylor Coll. of Med., Houston, TX; Cancer Genomics Program, Texas Children’s Hospital, Houston, TX; Department of Genetics, University of North Carolina at Chapel Hill, Chapel Hill, NC; Department of Pediatrics, University of California, San Francisco, San Francisco CA; Department of Pathology and Immunology, Washington University, St. Louis, MO; Division of Epidemiology, Department of Public Health Sciences, University of California, Davis, School of Medicine, Davis, CA; Population Sciences and Health Disparities Program, UC Davis Comprehensive Cancer Center, UC Davis Medical Center, Sacramento, CA; The Institute for Genomic Health, Icahn School of Medicine at Mount Sinai, New York, NY; Center for Translational Genomics, Icahn School of Medicine, New York, NY 10027; Division of Genomic Medicine, Department of Medicine, Icahn School of Medicine, New York, NY 10027; Department of Genetics and Genomic Sciences, Icahn School of Medicine, New York, NY 10027; Department of Epidemiology and Biostatistics, University of California San Francisco, San Francisco, CA

## Abstract

**Purpose:** Diagnostic yield from exome and genome sequencing varies widely across studies. It remains unclear how much of this variation reflects patient-level factors (e.g., sex, clinical features, race/ethnicity, genetic ancestry) versus site-level practices such as sequencing modality or variant interpretation workflows. We aimed to quantify the contributions of these factors to diagnostic outcomes across five U.S. clinical sequencing sites.

**Methods:** We performed a cross-sectional analysis of 3,008 prenatal, neonatal, and pediatric cases from the NHGRI Clinical Sequencing Evidence-Generating Research (CSER) consortium (2017–2023). Clinical indications spanned neurodevelopmental, neurological, immunological, metabolic, craniofacial, skeletal, cardiac, prenatal, and oncologic presentations. Genetic ancestry was inferred from sequencing data, and variants were interpreted using ACMG/AMP guidelines to classify DNA-based diagnoses. Generalized linear mixed models were used to estimate associations between diagnostic yield and fixed effects (sex, prenatal status, isolated cancer, number of clinical indications, sequencing modality, race/ethnicity, and genetic ancestry), while modeling study site as a random effect to quantify between-site variation.

**Results:** The overall diagnostic yield was 19.0%. Multiple clinical indications (OR=1.47, 95% CI 1.20–1.80, p<0.001) were associated with higher diagnostic yield, and male sex (OR=0.80, 95% CI 0.66–0.96, p=0.017) and prenatal status (OR=0.63, 95% CI 0.44–0.90, p=0.012) were associated with lower yield. Sequencing modality, race/ethnicity, genetic ancestry, and isolated cancer were not statistically significantly associated with diagnostic outcomes.. A model without fixed effects attributed ∼10% of variance in diagnostic yield to between-site differences. After adjusting for covariates, site-level variance decreased to 5.7%, indicating consistent variation across sites not explained by measured patient factors.

**Conclusion:** Across five sites, patient-level clinical features influenced diagnostic yield, but substantial site-level variation remained even after adjustment. Differences in variant interpretation, or case-classification practices may contribute to this residual variability. Further efforts to increase consistency in exome- and genome-sequencing diagnostic workflows may help reduce inter-site differences.

## Introduction

Genome and exome sequencing (henceforth sequencing) have become increasingly utilized tools for the genetic diagnosis of Mendelian conditions for a variety of clinical indications, such as neurodevelopmental disorders^1^, developmental delay^2^, multiple congenital anomalies^3^, intellectual disabilities^4,5^ and childhood neoplasms^6^. Sequencing has been used in a variety of clinical settings and has become the first-line tests for some clinical indications^3,5,7,8^, yet reported diagnostic yields vary widely across studies^9^. This variation may reflect differences in clinical populations, sequencing modalities, analytic workflows, and patient-level characteristics such as age, sex, and type or number of clinical indications⁹. Patient-level characteristics, including genetic ancestry, race/ethnicity, sex, and the number of clinical indications, are thought to contribute to diagnostic yield, yet how these factors compare with site-specific differences has not been systematically evaluated. Prior work has shown that laboratory-specific practices can also influence variant interpretation. For example, Amendola et al. compared variant classifications across multiple clinical laboratories using the ACMG/AMP framework and found substantial inter-laboratory discordance even when applying standardized guidelines^10^. However, that study did not include clinical information, leaving open whether site-level variation in diagnostic yield reflects analytic differences, patient mix, or both.

To address this, we analyzed 3,008 unrelated pediatric, NICU, and prenatal participants from five U.S. sites in the NHGRI CSER Consortium. Each site recruited patients and collected data independently, and these datasets were subsequently harmonized for cross-consortium analyses^11^. We evaluated diagnostic yield in relation to demographic and clinical covariates, genetic ancestry, and race/ethnicity while accounting for inter-site heterogeneity. This approach allowed us to assess how much of the variation in diagnostic yield reflects patient characteristics versus site-level differences, in a large, ancestrally diverse sample. The focus on diverse and underrepresented populations and harmonization across five sites provided a unique opportunity to address these questions – and in particular, whether sequencing yield is comparable in underrepresented and underserved populations.

## Methods

### Study populations and eligibility criteria

Participants were enrolled across five CSER studies at centers across the US between 2017 and 2022, each with the shared goal of evaluating the utility of diagnostic genomic sequencing in diverse and medically underserved individuals^11^. The studies included in the current analysis are: Texas KidsCanSeq at Baylor College of Medicine in Houston, TX^12^, NCGENES 2 at the University of North Carolina at Chapel Hill ^13^, NYCKidSeq at the Icahn School of Medicine in New York City, NY^14^, P^3^EGS at the University of California San Francisco^15^, and SouthSeq at the HudsonAlpha Institute for Biotechnology in Huntsville, AL^16^. All five studies enrolled children with a variety of clinical indications, including neurodevelopmental, neurological/neuromuscular, immunological, metabolic, craniofacial, skeletal, and cardiac phenotypes, as well as those with pediatric cancer diagnoses. One study site (P^3^EGS) also recruited prospective parents whose fetus had at least one structural anomaly detected on ultrasound^15^. Amendola, et al.^11^ describe the populations included, diversity targets for recruitment, locations and characteristics of the recruitment sites, medical conditions included, and study inclusion/exclusion criteria. Additional CSER publications^12–16^ provide study-specific information about IRB approvals and study protocols for each study. The participants were consented for data sharing and research analysis. Written informed consent was provided by adult participants ≥18 years of age, or by parents or legal guardians on behalf of their children <18 years of age or ≥18 years of age who were unable to consent independently. Assent was obtained from minors and intellectually disabled adults whenever possible. The current analysis complied with all relevant ethical regulations, including the Declaration of Helsinki.

In three of the five studies (KidsCanSeq, NCGENES 2, and SouthSeq), at least one parent voluntarily reported the race/ethnicity of their affected child as well as their own self-identified race/ethnicity. One study (NYCKidSeq) reported self-identified race/ethnicity of both parents and their affected child. In P^3^EGS, parents only reported their own race/ethnicity. To include P^3^EGS data in this study, we imputed race/ethnicity of the child or fetus based on the parent self-reported information (see Supplementary Methods). The parent and parent reported race and ethnicity data were standardized across all five studies^17^. One or more of the following categories were included for race/ethnicity characterizations: 1) “American Indian, Native American, Alaska Native”, 2) “Asian”, 3) “Black or African American”, 4) “White or European American”, 5) “Middle Eastern or North African/ Mediterranean”, 6) “Hispanic/Latino(a)”, 7) “Native Hawaiian/Pacific Islander”, 8) “Prefer not to answer”, 9) “Unknown/none of these fully describe me”.

The parent-reported race/ethnicity and sequence data were consented for data sharing through AnVIL (https://anvilproject.org/consortia/cser/resources).

### Genome and exome sequencing

All studies conducted genome or exome sequencing of affected probands (children or fetuses) and one study (P^3^EGS) also conducted exome sequencing of available biological parents. More details regarding methods and protocols used in sequencing and variant calling and annotation in the respective studies are provided elsewhere^12–16^ and in Supplementary Methods.

### Diagnostic outcomes/case classifications

Potential diagnostic variants were assessed by each study and reported using the ACMG/AMP variant classification framework and criteria^18^ as pathogenic (P), likely pathogenic (LP), or variant of uncertain significance (VUS). Harmonized case-level diagnoses were then made by each study based on variant pathogenicity and mode of inheritance criteria, as described in Supplementary Methods; cases were characterized as either positive (definitive positive, probable positive) and hence diagnosed, versus inconclusive or negative, and hence undiagnosed. Autosomal recessive designation required biallelic (homozygous or compound heterozygous) inheritance.

### Genetic ancestry, admixture, relatedness, and consanguinity estimation from sequence data

Principal component analysis (PCA) was performed on the 3,015 cases using the SMARTPCA program, part of the EIGENSOFT4.2 software package^19^. Individual genetic ancestry proportions were estimated using the ADMIXTURE software package ^20^ using a common set of 48,337 exome-wide markers. Genetic relatedness was estimated by kinship coefficients between all possible pairs of the 3015 cases using PC-Relate^21^. Further details are provided in Supplementary Methods.

### Statistical analyses

Diagnostic yield was modeled as a binary outcome (diagnosed versus undiagnosed) using generalized linear mixed models (GLMMs) with a logit link. All models included study site as a random intercept to account for clustering of individuals within sites, thereby allowing estimation of both within-site and between-site variance. Fixed effects included sex, number of clinical indications, prenatal status, sequencing modality (genome vs. exome), race/ethnicity categories, isolated cancer, and continuous genetic ancestry proportions (African, East Asian, Native American, Middle Eastern, and South Asian; European ancestry served as the reference).

We first fit a null model containing only the random intercept for site to quantify the proportion of variance in diagnostic yield attributable to site-level heterogeneity. We then fit a full model including all covariates as fixed effects and compared it to the null model. Statistical significance of the site-level random effect was assessed using likelihood ratio tests (LRT) with parametric bootstrap resampling.

To evaluate how different sources contributed to variability in diagnostic yield, we used variance decomposition of the GLMMs following the latent variable framework for logistic mixed models.^22,23^ Specifically, we summarized: Residual variance, representing individual-level variability not explained by the model; site variance, representing variance attributable to differences between sites; and fixed effects variance, representing the variation in diagnostic yield explained by the patient-level and clinical covariates included in the model. Odds ratios (ORs) and 95% confidence intervals (CIs) for fixed effects were obtained by exponentiating GLMM parameter estimates (Significance threshold: p<0.05).

We further assessed variance reduction in site-level differences stepwise, by sequentially adding covariates (e.g., ancestry, sex, clinical indications) to the null model and calculating the percentage reduction in site variance relative to the null.

All analyses were conducted in R (version 4.5.1) using the *lme4* package for GLMMs and the *performance* package for model diagnostics and variance decomposition. The mathematical framework for variance decomposition was as follows:

For each patient i in site j, diagnostic yield was modeled as:

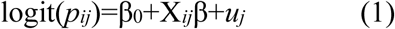

where *p_ij_* is the probability of a diagnosis for individual *i* at site *j*, β_0_ is the overall intercept, X*_ij_* is a vector of covariates with coefficients β, and *u_j_*∼*N*(0,σ^2^_site_) is the random intercept for site *j*. In logistic mixed models, the individual-level residual variance is fixed at π^2^/3≈3.29, reflecting the variance of the logistic distribution. The total variance (V_total_) can therefore be written as:

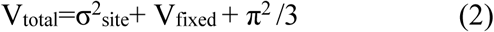

where V_fixed_ = Var(X^_ij_β^) is the variance of the linear predictor from fixed effects only, approximated from the fitted model. In the null model, V_fixed_ = 0, and equation (2) reduces to V_total_ = σ²_site_ + π²/3. Variance components were expressed as: residual variance π²/3, reflecting unexplained individual-level variability on the latent logit scale; site variance σ²site, representing between-site heterogeneity; and fixed effects variance V_fixed_, representing variation in diagnostic yield explained by the patient-level and clinical covariates included in the model. The proportional reduction in site-level variance attributable to covariates was calculated as:

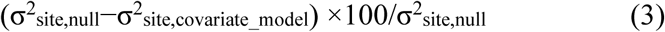

where σ^2^_site, null_ is the estimated site variance in the null model and σ^2^ _site, covariate_model_ is the site variance after including covariates. A detailed mathematical derivation of the variance decomposition is provided in the Supplementary Methods.

Chi-square tests were used to assess differences in variant types across study sites.

## Results

### Study population, and cohort characteristics

A total of 3,015 prenatal, NICU or pediatric cases from the five CSER studies had available sequencing data and clinical test outcomes and were included in this analysis. The number and description of cases from each of the five studies is provided in Table 1. The age range of the cases at study entry was 0 to 25 years old, with averages and standard deviations (SD) ranging from 24.4 (SD=5.61) weeks gestational age (GA) among the P^3^EGS prenatal cases; 0.98 (SD=1.78) months for the SouthSeq NICU cases; and 8.31 (SD=5.69) years for the 2,150 pediatric cases with age information across all studies. For the number of clinical indications per case, the overall ratio of more than one (>1) to one (1) was 0.7:1, and this ratio varied from 0.2:1 in KidsCanSeq which only required a cancer diagnosis for entry to 2.1:1 in NCGENES 2.

**Table 1.**
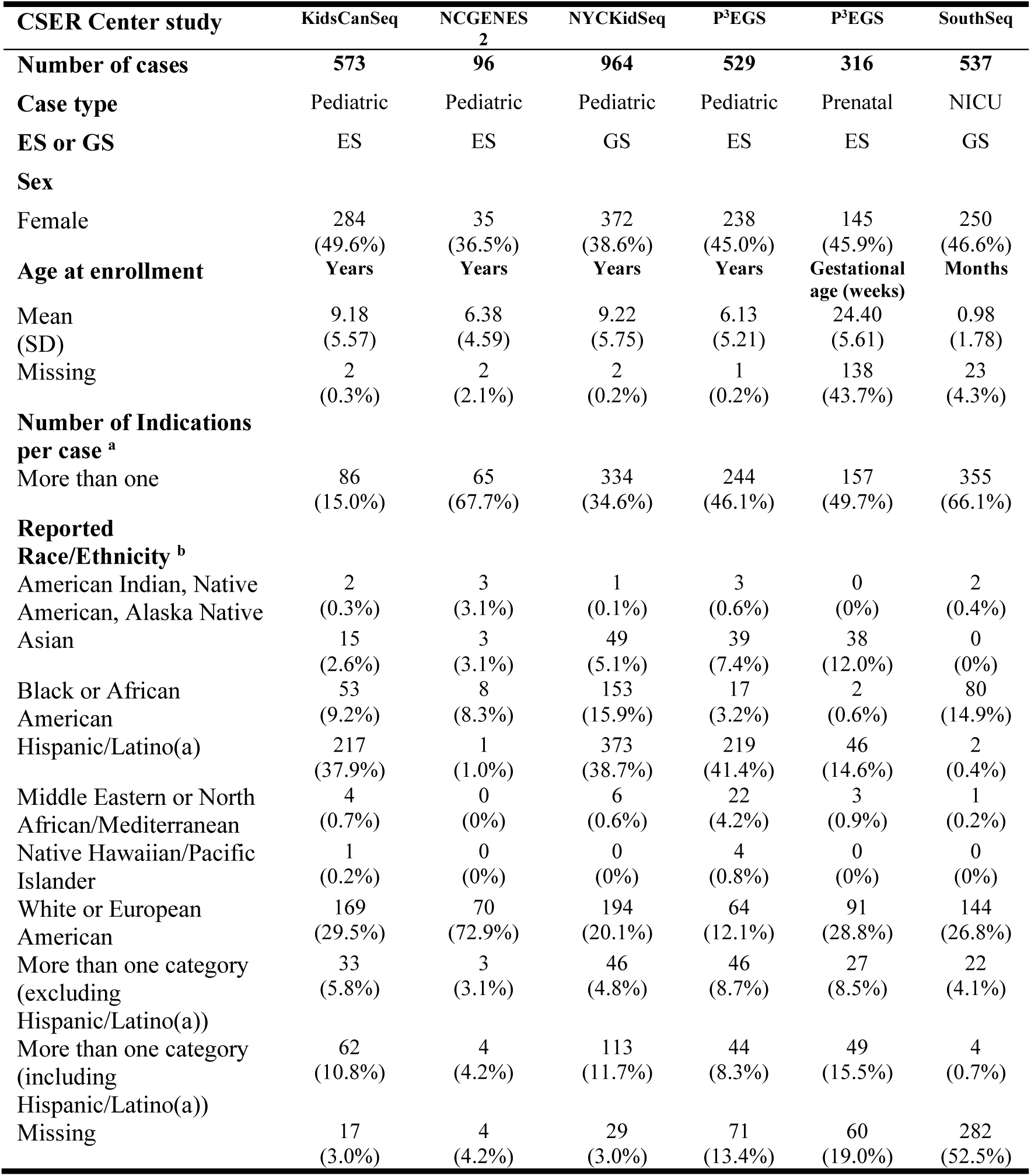
Description of the five CSER study cases.

Overall, the distribution of race/ethnicity of cases as reported by parents (or imputed in the case of P^3^EGS – see: Supplementary Methods) were: 28.46% Hispanic/Latino, 24.28% White/European American, 10.38% Black or African American, 4.78% Asian, 1.19% Middle Eastern or North African/Mediterranean, 0.36% American Indian, Native American, Alaska Native, 0.17% Native Hawaiian/Pacific Islander, and 15.02% multi-racial/ethnic. The remaining 15.36% were missing race/ethnicity information (i.e., “not reported”), or selected either “prefer not to answer” or “unknown/none of the categories fully describes me”. Supplementary Table 1 provides parents’ self-reported race/ethnicity, by study. Racial and ethnic representation differed across the studies (Table 1), reflecting both US regional differences and potentially divergent recruitment strategies.

The population genetic structure of the CSER cases, given by PC1 to PC6 of the PCA, with the Human Genome Diversity Program samples projected for interpretation, also shows an ancestrally diverse cohort (Supplementary Figures 1-3). Further details are given in Supplementary Results.

Individual genetic ancestry proportions of the 3015 cases, including those with unknown or non-reported race and ethnicity information, revealed some differences between the studies (Supplementary Figure 4). The means (SDs) of genetic ancestry proportions for the entire sample, reported in decreasing order, were European: 46.6% (33.8%), African: 19.4% (30.5%), Native American: 15.5% (24.7%), Middle Eastern: 9.3% (13.9%), East Asian: 4.5% (17.6%), South Asian: 4.3% (15.5%). The average estimated Oceanian genetic ancestry was 0.4% (1.2%) and was < 1% in all studies, so this category was not included in subsequent analyses due to low power. The correspondence between self-reported race/ethnicity and genetic ancestry is shown in Supplementary Figure 4, Supplementary Tables 2-8. Further details of study-specific genetic ancestry are given in the Supplementary Results.

### Diagnostic yield by CSER study

Seven pairs of full siblings were identified and one of each pair removed, leaving 3,008 unrelated cases for all subsequent analyses. (see Supplementary Methods for relatedness analysis) across the five studies, 573 (19.0%) had a diagnostic case outcome. The diagnostic yield varied across studies (8.2%, 7.3%, 17.3%, 23.8%, and 28.0% for KidsCanSeq, NCGENES 2, NYCKidSeq, P^3^EGS and SouthSeq, respectively).

Overall, the diagnostic yield was higher in female than male cases (20.7% vs 17.1%), as was also true within each study (Supplementary Table 9). The diagnostic yield in cases who had more than one clinical indication was higher than in cases with a single indication (24.1% vs 15.0%) as was also true within each study (Supplementary Table 9). On average, studies that used genome sequencing had a higher overall diagnostic yield compared to studies that used exome sequencing (21.2% vs 16.9%), but comparison within studies was impossible because no study used both, so this comparison is confounded by other study-specific differences. By contrast, we observed no difference in diagnostic yield across the various race/ethnicity groups, as follows: 18.6% Hispanic/Latino, 17.2% White/European American, 16.6% Black or African American, 16.8% Asian, 19.4% Middle Eastern or North African/Mediterranean, 27.3% American Indian, Native American, Alaska Native, 18.6% More than one category (excluding Hispanic/Latino(a)), 16.3% More than one category (including Hispanic/Latino(a)), and 27.4% Missing. Apart from those with missing information (majority from SouthSeq), the other race/ethnicity categories did not differ significantly in diagnostic yield from White/European American overall (p >0.05, Fisher’s Exact test). The rate of missingness in SouthSeq data reported here contrasts with the race/ethnicity data reported in Bowling et. al 2022^16^ as this analysis used only self-reported data, while Bowling et. al 2022 included clinician-reported demographics.

**Figure 1:**
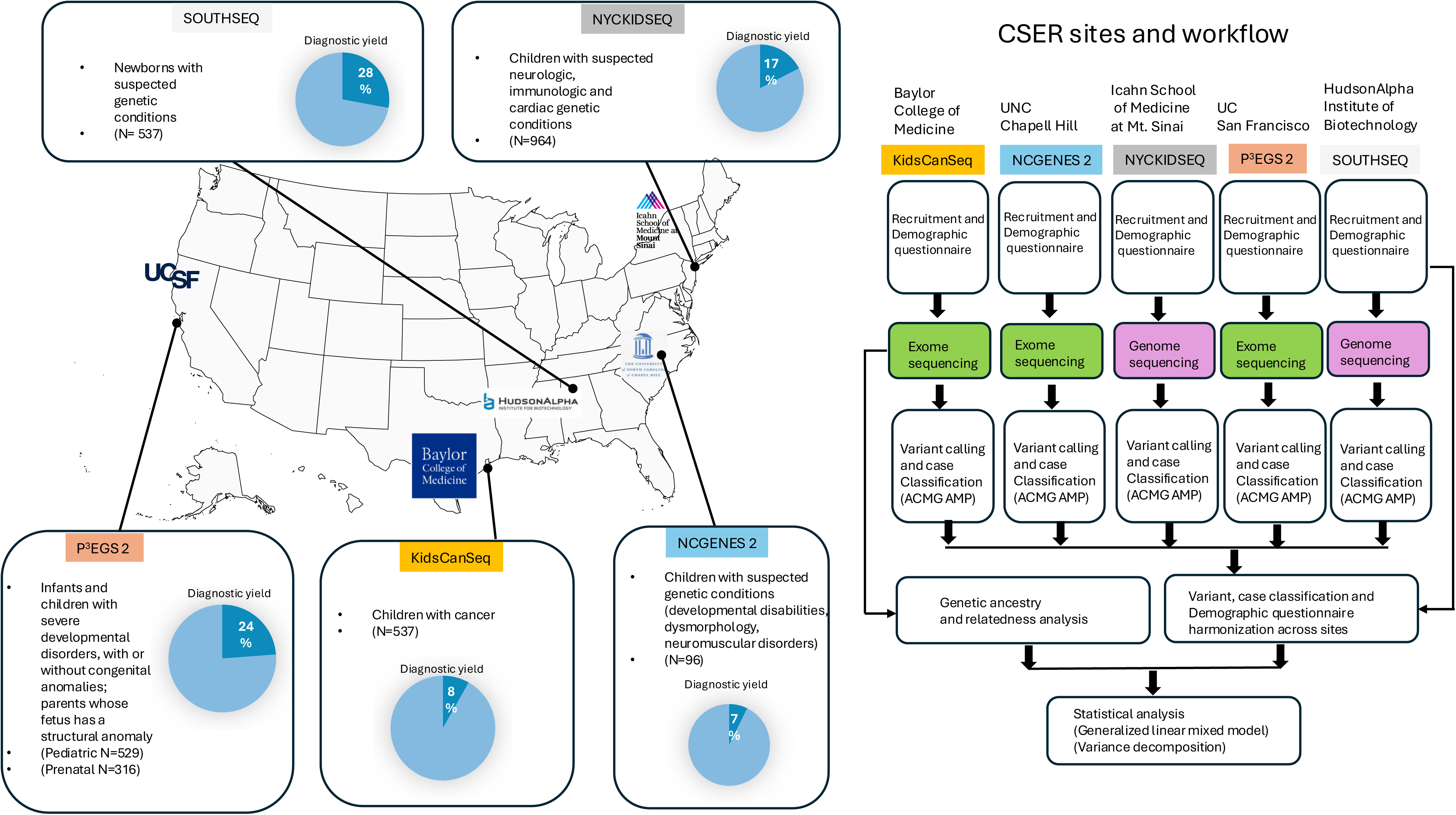
Schematic of CSER sites, inclusion criteria, processing flowchart and diagnostic yield.

Across all studies, most positive cases (75.9%) had single P/LP variants in genes for autosomal dominant disorders, ranging from 68.2% in P^3^EGS to 93.6% in KidsCanSeq (Supplementary Table 10). Homozygous or compound heterozygous P/LP variants in genes with autosomal recessive inheritance accounted for 13.3% of positive cases total, ranging from 2.1% in KidsCanSeq to 19.9% in P^3^EGS (Supplementary Table 10). X-linked mode of inheritance accounted for 8.9% of positive cases total, ranging from 4.3% in KidsCanSeq to 14.3% in NCGENES 2 (Supplementary Table 10).

### Mode of Inheritance and Consanguinity

Among autosomal recessive cases, homozygous variants were more frequent in studies with greater parental relatedness (P^3^EGS and SouthSeq). Consanguinity coefficients (F) were generally close to zero across the cohort but were higher among positive cases with homozygous recessive inheritance. In P^3^EGS, mean F was significantly higher in homozygous AR cases compared with undiagnosed cases (difference = 0.044, p = 0.0004), with similar non-significant trends observed in NYCKidSeq and SouthSeq. Overall, 14.6% of unrelated individuals had F ≥ 0.0156, the expected value for offspring of second cousins. Higher F values were associated with higher Middle Eastern and South Asian ancestry among homozygous AR cases. Individuals with F ≥ 0.0156 had mean ancestries of 15.1% Middle Eastern and 30.0% South Asian, compared with 6.1% and 4.7%, respectively, among those with F < 0.0156 (Supplementary Results).

Ancestry-related consanguinity did not affect overall diagnostic yield. Although most diagnoses were autosomal dominant, homozygous recessive diagnoses associated with parental relatedness were present across studies.

### Variant types by Study

P/LP variant type frequencies differed significantly across studies (χ² = 19.15, p = 0.024; Supplementary Table 16). Frameshift variants were most common in KidsCanSeq, while missense variants predominated in NYCKidSeq, P^3^EGS, and SouthSeq. Splice-site variants were more frequent in KidsCanSeq and SouthSeq, and CNVs accounted for 28–29% of P/LP variants in NYCKidSeq and SouthSeq but were rare (<1%) elsewhere. Most P/LP variants (567/575) had ancestry-specific allele frequencies <0.001 in gnomAD v4 or were absent entirely. These inter-study differences in variant type frequencies likely reflect variation in clinical indications, sequencing methods, and annotation practices.

Further details on genetic ancestry, consanguinity, modes of inheritance including autosomal recessive inheritance are provided in the Supplementary Results (Supplementary Figure 5-8, Supplementary Tables 10-15), along with details regarding P/LP variant types, population frequencies, and recurrence (Supplementary Tables 16-18).

### Associations of Demographic and Clinical Covariates with Diagnostic Yield

In multivariable generalized linear mixed models (GLMMs) that included site as a random effect across all 3,008 cases, several patient-level and clinical fixed effects were significantly associated with diagnostic yield. Males had lower odds of receiving a diagnosis compared to females (OR = 0.80, 95% CI 0.66–0.96, *p* = 0.017). Individuals with two or more clinical indications had higher odds of a positive result relative to those with a single indication (OR = 1.47, 95% CI 1.20–1.80, *p* < 0.001). Prenatal cases had lower odds of receiving a diagnosis compared with non-prenatal cases (OR = 0.63, 95% CI 0.44–0.90, *p* = 0.012) (Figure 2).

**Figure 2:**
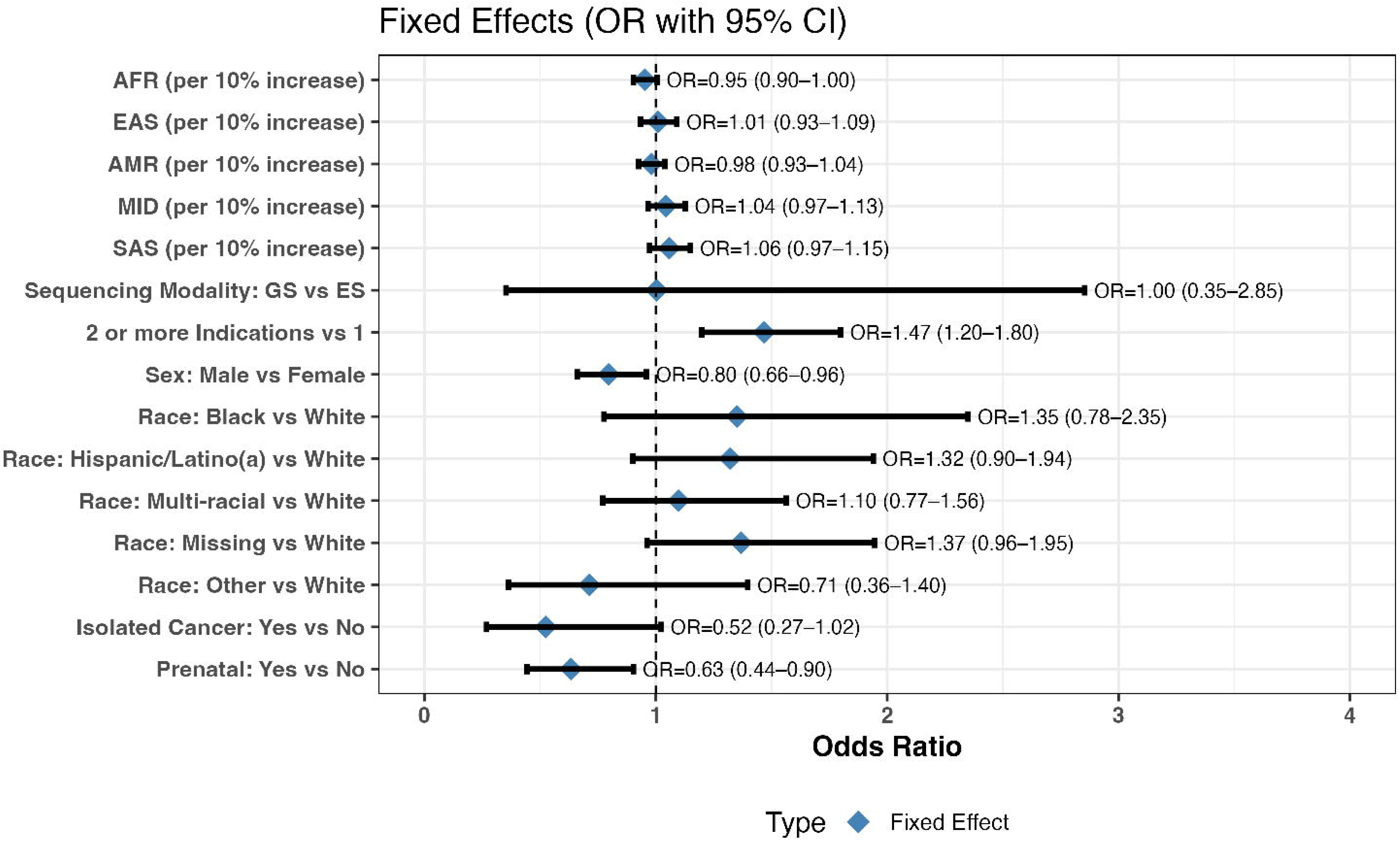
Association of Study Factors with Diagnostic Yield (GLMM Fixed Effects) Forest plot showing odds ratios (ORs) and 95% confidence intervals for fixed effects from the full generalized linear mixed model. Covariates include genetic ancestry (per 10% increase; European ancestry as reference), demographic and clinical characteristics, and sequencing modality, with adjustment for study site as a random effect. Odds ratios greater than 1 indicate higher odds of a positive diagnostic yield.

Sequencing modality (genome vs exome; OR = 1.00, 95% CI 0.35–2.85, p = 0.995) and race/ethnicity categories were not significantly associated with diagnostic yield. Isolated cancer showed a trend toward lower diagnostic yield (OR = 0.52, 95% CI 0.27–1.02, p ≈ 0.054) that did not reach statistical significance, possibly reflecting limited power due to the small number of isolated cancer cases across sites (Figure 2).

Genetic ancestry (modeled as continuous predictors) was not significantly associated with diagnostic yield for any ancestry group. Odds ratios per 10% increase in ancestry proportion were close to 1.0 for: African (OR = 0.95, 95% CI 0.90–1.00), East Asian (OR = 1.01, 95% CI 0.93–1.09), Native American (OR = 0.98, 95% CI 0.93–1.04), Middle Eastern (OR = 1.04, 95% CI 0.97–1.13), or South Asian (OR = 1.06, 95% CI 0.97–1.15) ancestries, with European ancestry as the reference ancestry (Figure 2).

### Inter-Site Heterogeneity in Diagnostic Yield

Site-level differences in diagnostic yield were statistically significant. Including study site as a random effect in the generalized linear mixed model substantially improved model fit (LR = 81.55, p < 0.001; parametric bootstrap p < 0.001). In the null model, 10.1% of the variance in diagnostic yield was attributable to differences between sites. After accounting for all patient-level and clinical covariates, site-level variance decreased to 5.7% — meaning the measured covariates explained approximately 43% of the between-site differences, leaving 57% unexplained (Table 2).

**Table 2:** Sources of variation in diagnostic yield across sites.

| **Model** | **Residual**<br>**(Case-to-case variability)**<br>**(%)** | **Differences between sites**<br>**(%)** | **Fixed**<br>**effects**<br>**(%)** |
| --- | --- | --- | --- |
| No Fixed effects eg sex<br>(Null) | 89.9 | 10.1 | 0.0 |
| Full model (with all fixed<br>effects) | 90.0 | 5.7 | 4.3 |

When covariates were added individually, isolated cancer and prenatal status had the strongest effects on reducing between-site variance (54% and 44% reductions respectively). Although isolated cancer was not associated with diagnostic yield at the individual level, including it reduced between-site variance, suggesting that sites differed in the proportion of isolated cancer referrals. Other covariates, ancestry, sex, sequencing modality, and race/ethnicity, had smaller effects. Overall, patient-level factors explained a moderate portion of why sites differed, but substantial between-site variation remained.

## Discussion

The CSER program focused on evaluating clinical utility of exome and genome sequencing in a diverse sample of individuals from underrepresented and underserved backgrounds, to fill the gaps in knowledge of its utility in these groups. Prior studies from this program have examined clinical utility in terms of post-diagnostic care^24^, as well as participant inclusion and diversity^25–29^. The focus of this study was to assess associations of clinical and demographic covariates with diagnostic yield and the contribution of both individual-level covariates and study site variability in diagnostic outcomes.

Among 3,008 cases with suspected Mendelian disorders from five CSER studies, we observed no association between genetic ancestry, self-reported race/ethnicity or sequencing modality and diagnostic yield. Sex (males with lower odds), number of clinical indications (single indications with lower odds), and prenatal status (lower odds) were significant predictors of diagnostic yield. Isolated cancer (lower odds) was not statistically significant at the individual level but did contribute to explaining between-site differences. Overall, these covariates accounted for a moderate portion (43%) of the among site variance.

Some of these findings contrast with prior studies. For example, regarding ancestry, the DDD study^30^ reported reduced diagnostic yield in individuals of African or Middle Eastern ancestry. In DDD, lower yield was attributed to both lack of ancestry-matched control allele frequencies and the higher likelihood of African ancestry individuals being sequenced as singletons rather than trios, which reduced detection of pathogenic variants. Our analysis included five studies with different sequencing strategies. Four of the studies sequenced probands initially, followed by Sanger sequencing of probands and parents for validation and inheritance determination. One study (P^3^EGS) performed trio and duo sequencing, as well as a “proband-first” approach^15^. In that study, diagnostic yield did not differ significantly by the number of parents sequenced, contrasting with the 4.7-fold lower yield observed for singletons in DDD. Differences in parental participation likely reflect social factors linked to self-reported race/ethnicity rather than genetic ancestry, although the two are sometimes correlated. In our study, potential P/LP variants were not filtered using ancestry-matched allele frequencies; instead, variants were classified using ACMG/AMP criteria for pathogenicity. This standardized approach likely mitigates ancestry-related biases, highlighting that when robust variant interpretation frameworks are applied, diagnostic yield does not differ by genetic ancestry. Furthermore, diagnostic yield did not differ by self-reported race/ethnicity, suggesting that social factors correlated with race/ethnicity also did not impact diagnostic yield in our setting. Differences between our findings and DDD may also reflect study-specific factors, such as clinical severity or variant annotation pipelines^31^.

Site-level heterogeneity emerged as an additional source of variation in diagnostic yield, even after accounting for all the clinical and demographic covariates. There are several possibilities for residual site differences contributing to this heterogeneity. On the clinical side, in terms of clinical severity, we only had harmonized information on whether there was a single indication for testing or more than one. Thus, it is possible this variable did not capture all differences in severity across sites. For example, SouthSeq, which had the highest diagnostic yield, included infants in the NICU, who may have had a greater level of severity than other sites recruiting pediatric participants who were generally older. On the other hand, phenotypic assessment instruments were coordinated across sites through working groups in an attempt at harmonization^11^. Recruitment numbers and types of individuals included did vary across sites, from prenatal to NICU to pediatric cases varying in age. Recruitment across sites was also impacted by the Covid-19 pandemic, which differentially curtailed recruitment at sites (e.g. some sites enabled telehealth recruiting while others did not). While impact on recruitment influenced the total number of individuals recruited per site, there is no indication it impacted the diagnostic yield of those that were recruited.

Sites did also differ in terms of sequencing modality, with two sites employing genome sequencing and three sites exome sequencing. In addition, the sites using genome sequencing did not require probands to have a prior negative array (a test for diagnostic CNVs) whereas the exome sequencing sites did. This also manifested in the two sites employing genome sequencing having a higher rate of diagnostic CNVs. However, sequencing modality was not a significant predictor of diagnostic yield in our GLMM analysis. While SouthSeq, which had the highest diagnostic yield, used genome sequencing, the diagnostic yield of NYCKidSeq, which also used genome sequencing, was lower than the diagnostic yield of P^3^EGS, which used exome sequencing.

Differences in mode of inheritance and consanguinity may also have contributed to variation across studies. While most diagnoses were autosomal dominant, cohorts differed in the proportion of autosomal recessive cases, which were more frequent in studies with greater parental relatedness. Consanguinity was significantly associated with homozygous recessive diagnoses in P^3^EGS and trended higher in NYCKidSeq and SouthSeq, consistent with prior reports that increased parental relatedness modestly elevates diagnostic yield for recessive conditions^32^.

Another possibility underlying site-specific variability is differences across sites in variant annotations and diagnostic calls. The CSER consortium made an effort to harmonize many aspects of the program, including variant annotation and case-level diagnoses. This included a Sequencing and Diagnostic Yield (SADY) working group which met routinely to discuss harmonization of annotation and diagnostic practices^10^. All sites used ACMG-AMP criteria for variant annotation, and employed expert evaluation teams including molecular pathologists, clinical geneticists, genetic counselors, statistical geneticists, informaticians and bioethicists.

One project of this working group was to compare annotation practices across the 6 CSER sites plus 3 other molecular laboratories (9 sites total) for a set of 158 variants considered for secondary/incidental findings^10^. In that study, variants submitted by one lab were independently annotated by two of the others, and discrepancies evaluated. There were no extreme disparities for variants initially annotated as P or LP to B or LB or initially annotated as B or LB to P or LP; however, some P and LP variants were classified as VUS by the independent labs. Specifically, among 27 variants initially annotated as P or LP, 9 (33%) were annotated as VUS by the independent labs. Subsequent discussions occurred among the group of the discordant variants, and after adjudication the number of P or LP variants independently classified as VUS was reduced to 5 (19%). Thus, differences in variant classifications that potentially impacted diagnoses did occur among the groups. This is likely because of incomplete clarity and differing interpretations regarding implementation of some of the ACMG-AMP guidelines^10^. It should also be noted that this study did not assess if there were significant overall systematic differences across labs in their variant calling practices (e.g. if some were more conservative than others or the impact of different variant calling or interpretation pipelines). However, it did show that broader discussions among the labs did reduce the differences observed from 33% to 19%.

ACMG-AMP criteria are focused on variant interpretation (e.g. as P, LP, VUS, LB or B). However, diagnostic yield is based on case-level diagnoses, which invokes other criteria beyond simple variant annotation, for example including the mode of inheritance, the combination of variant interpretations and phase for compound heterozygous variants, whether variants are inherited or de novo, and the degree of clinical overlap between the phenotype(s) of the tested individual and what is paradigmatic for the identified gene and variant. For the latter there are no specific criteria regarding the degree of overlap, and it remains a subjective decision based on the knowledge and experience of the clinical geneticists and genetic counselors who assess the patients. Thus, one can expect all these factors to play a potential role in variation across sites in diagnostic yield.

A study similar to the variant annotation study mentioned^10^ based on case-level diagnostic practices across different labs would help illuminate the reasons for the residual DY variation we observed that also likely exists across other groups. However, some challenges may exist for such a study that need to be overcome, including the sharing of PHI-protected clinical data across sites.

In any event, while we cannot assert direct evidence that site-specific variant calling, variant annotation/interpretation and case-level diagnostic practices explain the residual variance we observed across the CSER sites, the indirect evidence mentioned above remains a plausible explanation. It is simply a reflection of the fact that while there are guidelines for variant annotation, there is still room for differences in how those guidelines are implemented for diagnoses, and under current practices differences in diagnostic rates across well-trained sites is likely inevitable.

In terms of generalization to other testing groups, we believe the factors as described above that led to differences across our 5 CSER sites likely contribute to variability across all testing entities. In fact, because of CSER’s attempt at harmonization of various aspects of clinical sequencing, the variability we observed across the CSER sites might actually be less in comparison to broad variability across centers observed more generally^9^.

## Conclusion

In summary, this multi-site analysis of pediatric, NICU, and prenatal cases shows that diagnostic yield from exome and genome sequencing can vary due to a number of clinical and technological factors. Among individual-level covariates, sex and clinical severity (number of clinical indications) were significant predictors, along with prenatal status. On the other hand, race/ethnicity and genetic ancestry and sequencing modality had minimal influence. Accounting for all these clinical, demographic and technical variables explained 43% of the variation in diagnostic yield across sites, leaving a significant 57% unaccounted for, potentially due to site differences in variant annotation and case-level diagnostic practices. Further efforts expanding on current diagnostic guidelines, in particular refinement of variant annotation and assessment of clinical fit of a tested individual’s phenotype with those characteristic of a particular gene and its variants should enable greater consistency of exome and genome sequencing-based diagnosis.

## Data Availability

The datasets used in this study are available on AnVIL: https://anvilproject.org/consortia/cser/resources.

Access to CSER data is moderated through dbGaP data access requests. The following phsIDs are associated with CSER consortium data: phs001089. v3.p1 (SouthSeq), phs002324.v1.p1 (P3 EGS), phs002337.v1.p1 (NYCKidSeq), phs002110.v1.p1 (NCGENES 2), phs002378.v1.p1 (KidsCanSeq).

## Code Availability

The code supporting the analyses is available from the corresponding author upon request.

## Acknowledgements

This work was funded by: U01 HG006485, U01 HG007301, U01 HG006487, U01 HG009610, U01 HG009599, U24 HG007307

## Author Contributions

Conceptualization: N.R., Y.M.

Data curation: Y.M., D.C., K.F., J.M.L., S.K., M.L.T., B.C.P.

Formal analysis: Y.M., N.R.

Funding acquisition: L.H., G.P.J., B.A.K., P.K., M.N., S.E.P., B.C.P., A.S., E.E.K., N.R.

Investigation: Y.M., N.R. Methodology: Y.M., N.R.

Project administration: L.H., Y.M., N.R.

Resources: J.M.G., L.H., G.P.J., S.K., M.K., B.A.K., P.K., M.N., S.E.P., B.C.P., A.S., M.L.T., A.B.P., E.E.K.

Supervision: N.R., S.P. Visualization: Y.M.

Writing – original draft: Y.M.

Writing – review & editing: D.C., K.F., J.M.L., J.M.G., L.H., G.P.J., S.K., B.A.K., M.K., P.K.,

M.N., S.E.P., B.C.P., A.S., M.L.T., A.B.P., E.E.K., N.R.

## Declaration of AI and AI-assisted technologies in the writing process

During the preparation of this work, the authors used ChatGPT to rephrase certain sections to enhance readability and to assist with the article’s translation.

## Ethics Declaration

This study involved human participants and was conducted in accordance with all relevant ethical regulations, including the Declaration of Helsinki. Each participating CSER site obtained Institutional Review Board (IRB) approval prior to participant enrollment: SouthSeq was approved by the University of Alabama at Birmingham IRB (IRB-300000328); KidsCanSeq was approved by the Baylor College of Medicine IRB (protocol H-42376); and P^3^EGS was approved by the University of California San Francisco IRB (17-22504 and 17-22420), and the Fresno Community Medical Center IRB (protocol 2019024). NYCKidSeq was approved by the IRBs at the Icahn School of Medicine at Mount Sinai and Albert Einstein College of Medicine.

NCGENES 2 was approved by the Biomedical IRB of the University of North Carolina, Chapel Hill (IRB #17-0816). Written informed consent was provided by adult participants (≥18 years of age), or by parents or legal guardians on behalf of children (<18 years of age) or adults ≥18 years of age who were unable to consent independently. Assent was obtained from minors and adults with intellectual disabilities whenever possible. All participants consented to data sharing and research analysis.

