## Supplementary material for "Determinants of DNA-sequence-based Diagnostic Yield in the CSER Consortium"

### **Supplementary Methods**

#### **Imputation of race/ethnicity for cases/offspring in P^3^EGS**

Since P^3^EGS parents only reported their own race/ethnicity and not for their offspring (‘case’), we imputed race/ethnicity of P^3^EGS case offspring using the self-reported race/ethnicity of both parents of each of the cases. Parents were asked to respond to one or more of the following categories that best describe their race/ethnicity: 1) “American Indian, Native American, Alaska Native”, 2) “Asian”, 3) “Black or African American”, 4) “White or European American”, 5) “Middle Eastern or North African/ Mediterranean”, 6) “Hispanic/ Latino(a)”, 7) “Native Hawaiian/Pacific Islander”, 8) “Prefer not to answer”, 9) “Unknown/none of these fully describe me” (Select all that apply).

If both parents reported the same (single) R/E category, the child was assigned that R/E category. For example, if parent “a” reported “Asian”, and parent “b” reported “Asian”, then the child would be assigned “Asian”. This includes the categories “Prefer not to answer”, “Unknown/ none of these fully describe me”. If both parents did not report race/ethnicity, the child’s race/ethnicity would be imputed as “Not reported”.

If the two parents each reported a single R/E category, and they were discordant, the child was assigned “More than one category (excluding Hispanic/Latino(a)” if neither parent reported Hispanic/Latino(a) or “More than one category (including Hispanic/Latino(a))” if either parent did report Hispanic/Latino(a). If one or both parents reported more than one category, then the child was assigned “More than one category (excluding Hispanic/Latino(a)” or “More than one category (including Hispanic/Latino(a))” based on the logic stated above.

#### **Genome and exome sequencing**

Briefly, P^3^EGS, NCGENES 2, KidsCanSeq conducted exome sequencing, while NYCKidSeq and SouthSeq conducted genome sequencing. For P^3^EGS and KidsCanSeq, the DNA sequences were aligned to the human genome reference build GRch37/Hg19, while the rest were aligned to build GRch38/Hg38. To facilitate uniform statistical analysis of genome sequencing and exome sequencing across the studies, a set of 48,337 SNPs common to sequence data from all five studies was created for the reported diagnostic yield and genetic ancestry analyses.

#### **Derivation of SNPs for ancestry related analyses**

Variants in VCF files from NCGENES 2, NYCKidSeq, P^3^EGS, and SouthSeq with sequencing depth at or below 10 (DP<=10) and genotype quality equal to or less than 20 (GQ <=20) were filtered out using GATK^1^. VCF files from P^3^EGS and KidsCanSeq were then lifted over from human genome reference version GRCh37 to GRCh38 using the Picard tool in the GATK suite of tools^1^. Human Genome Diversity Panel (HGDP) GS samples from the gnomAD V3^2^ call set were used as the reference for genetic ancestry and admixture estimation (N=829 unrelated individuals). The HGDP samples were all mapped to the GRCh38 reference sequence. High-performance markers were selected from the intersection of HGDP and combined studies variant data for downstream genetic ancestry, admixture, relatedness, and consanguinity analysis using the following criteria:

1) Markers with minor allele frequency (MAF) >= 0.05 in any of 7 populations in gnomAD HGDP unrelated individuals: i) African (ii) Native American, iii) South Asian iv) East Asian, v) European, vi) Middle Eastern, vii) Oceanian, in the intersected sequenced (exome-wide) regions of samples from the five studies were retained (N=97,383 markers).

2) Restriction of markers in the gnomAD HGDP dataset from step 1 above by intersecting with the exome wide markers in the combined studies dataset. This was conducted using Bcftools^3^. A total of N=50,531 exome wide markers remained.

3) Only biallelic, autosomal SNPs, with a call rate > 95% (in either HGDP or individual study cohort data separately) were retained. Variants in regions known to affect principal components (PCs) (HLA region on chromosome 6p, inversion on chromosome 8p23 and inversion on chr 17q21, GRCh38 build) were removed^4^.

After all the above steps, 48,337 SNPs remained to be used for genetic ancestry, genetic admixture, genetic relatedness, and consanguinity analyses on the common set of cases.

#### **Principal Components Analysis**

After linkage disequilibrium (LD) pruning (0.5 kb in a 5000 kb window), 36,366 high-performance markers from an initial 48,337 marker set (see above) were used for the PCA. The Human Genome Diversity Panel (HGDP)^5^ reference sample genome sequence results obtained from gnomAD^2^ were then used to project those individuals onto the derived PCs to facilitate ancestral geographic interpretation of the cases^6^.

#### **Admixture Analyses**

We used a supervised approach, whereby unrelated individuals in (K=7) reference categories were assigned 100% genetic ancestry to the reference-labelled category, and individual genetic admixture proportions were estimated in 3,015 cases from the five studies. We used K=7 continental/sub-continental ancestral reference groups from the Human Genome Diversity Project (HGDP) panel^7^. The 7 reference categories were: African - Afr (Yoruba, Mandenka N=40), Native American – Amr (Colombia, Karitiana, Surui, Pima, N=40); East Asian - Eas (Han Chinese, Japanese N=40); Middle Eastern - Mid (Druze, Palestinian, Bedouin, N=40); European - Eur (French, Orcadian, Tuscan, Sardinian, N=40); Oceanian - Oce (Papuan, Melanesian N= 30); and South Asian - Sas (Pathan, Sindhi, N=39)^5^. The individual genetic admixture proportions were visualized using Pong software ^8^.

#### **Genetic Relatedness**

Genetic ancestry was controlled by using the first 8 PCs from PCA in PC-Relate to estimate only recent genetic relatedness due to family structure (HGDP reference samples were also used in the analysis as recommended). LD pruning (0.1 kb in a 10000 kb window) was done on the initial 48,337 markers to select a set of independent SNPs (N=36,484) for the relatedness analysis including the PCs used for control of ancestry. Similarly, consanguinity coefficients for cases were estimated using PC-relate, also controlling for ancestry using the first 8 PCs. The consanguinity coefficient (F) as used in this study is the probability that two alleles at a locus in an individual are identical by descent from a common ancestor but estimated by the self-kinship coefficient of the two alleles in an individual and can therefore be negative^9^. The expected consanguinity coefficient of children of first cousins is 1/16 (0.0625), and of second cousins is 1/64 (0.0156).

#### **Algorithm for Case-level Diagnoses**

All patients received a case-level diagnosis as either definitive positive, probable positive, inconclusive, or negative. The classification was developed by the Sequencing and Diagnostic Yield (SADY) Working Group of the CSER consortium as in the table below^10^:

Definitive Positive

- Implicated variant(s) are pathogenic
- Phenotype and inheritance pattern are consistent with patient’s condition
- Known phase or *de novo* status

Probable Positive

- Implicated variant(s) are likely pathogenic or a combination of pathogenic/likely pathogenic
- For recessive condition, combination of a pathogenic/likely pathogenic variant with a variant of uncertain significance provided no other inconclusive conditions (below) exist
- Phenotype and inheritance pattern are consistent with patient’s condition
- Known phase or *de novo* status, or for recessive condition with only one parent/sibling available, that family member has only one of the implicated variants

Inconclusive

Potential diagnostic variant(s) detected with one or more of the following contributions to case-level ambiguity:

- Unknown phase
- Variant of uncertain significance
- Insufficient zygosity (heterozygous variant in a recessive disease-associated gene)
- Insufficient phenotype match
- Novel gene

#### **Variance Decomposition and Random Effect Testing**

For individual i in site j we fit a logistic mixed model with a site random intercept:

logit(*p_ij_*)=β_0_+X*_ij_*β+*u_j_,u_j_*∼N(0,σ^2^_site_)

where:

p*_ij_*=Pr (Y*_ij_*=1) is the probability of a positive outcome, X*_ij_* is the covariate vector, β are fixed-effect coefficients, *u_j_* is the site-specific random intercept.

The variance of the standard logistic distribution is

σ^2^_resid_=π^2^ /3≈3.29

This represents the individual-level residual variance on the latent logit scale. **Site variance** is the estimated variance of the random intercept:

$\hat{\text{σ}}$^2^_site_=Var ($\hat{u}$*_j_*).

**Fixed-effects variance** is approximated by the variance of the predicted linear predictor excluding random effects:

V_fixed_=Var (X*_ij_*$\hat{\beta}$).

The total variance on the latent scale is

V_total_=σ^2^_site_+V_fixed_+σ^2^_resid_.

In the null model, V_fixed_ = 0, and V_total_ reduces to σ²_site_ + π²/3. This framework follows the latent variable approach of Nakagawa and Schielzeth (2013)^11^ and Nakagawa, Johnson and Schielzeth (2017)^12^, and corresponds to the marginal R² = V_fixed/_V_total_ and conditional R² = (V_fixed_ + σ²_site_)/V_total_.

The percent contributions are as follows:

**Residual %:**

(100×π^2^/3) /(σ^2^_site_+V_fixed_+π^2^/3)

**Site %:**

(100×σ^2^ _site_ )/(σ^2^_site_+V_fixed_+π^2^/3)

**Fixed %:**

(100×V_fixed_ )/(σ^2^_site_+V_fixed_+π^2^/3)

For the null model, V_fixed_=0.

The percent reduction in between-site variance when moving from the null to a covariate-adjusted model is

%Δσ^2^_site_=(100×σ^2^_site,null_−σ^2^_site,covariate_model_ )/σ^2^_site_,_null_.

**Likelihood ratio test (LRT):**

The likelihood ratio test (LRT) was used to evaluate whether including additional model terms, in this case, random effects, significantly improved model fit. Let $\theta_{\text{GLMM}}$ denote the parameter vector for the full generalized linear mixed model (GLMM), consisting of the fixed-effects coefficients $\beta$ and the random-effect variance components (e.g., the site-level variance $\sigma_{\text{site}}^{2}$). Let $\theta_{\text{GLM}}$ denote the corresponding parameter vector for the reduced generalized linear model (GLM) that excludes random effects and therefore contains only the fixed-effects parameters.

The observed LRT statistic is defined as

$$\Lambda_{\text{obs}}=2[\mathcal{l}(\hat{\theta}_{\text{GLMM}})-\mathcal{l}(\hat{\theta}_{\text{GLM}})],$$

where $\mathcal{l}(\cdot)$ is the maximized log-likelihood of the fitted model under the respective parameter estimates. Under standard regularity conditions, $\Lambda_{\text{obs}}$ follows a chi-squared distribution with degrees of freedom equal to the difference in the number of estimated parameters. However, when testing variance components (e.g., the null hypothesis $\sigma_{\text{site}}^{2}=0$), the null lies on the boundary of the parameter space because variances must be non-negative. This violates the usual asymptotic assumptions and renders the chi-squared approximation conservative.

To obtain valid p-values, we therefore used parametric bootstrapping with 1,000 simulated datasets, implemented using the **PBmodcomp** function from the *pbkrtest* R package. For each bootstrap sample, both the full and reduced models were refit, and the LRT statistic was computed. The resulting empirical distribution of LRT values was used to obtain a bootstrap-based p-value by comparing $\Lambda_{\text{obs}}$ to its simulated null distribution. This approach appropriately accounts for the hierarchical structure of the data and the boundary constraints inherent in variance-component testing^13^.

### **Supplementary Results**

#### **PC Results**

The first PC distinguishes recent African ancestry from all others while the second PC distinguishes European, Native American, and East Asian ancestries (Supplementary Figure 1). The third PC distinguishes Native American vs East Asian ancestry, while the fourth PC distinguishes European and South Asian ancestries (Supplementary Figure 2). The fifth PC identifies Middle Eastern ancestry (Supplementary Figure 3). Many CSER cases appear to form a continuum along and between the axes of PCs, except for clusters corresponding to individuals of European, East Asian, and South Asian genetic ancestries.

#### **Genetic Ancestry and correspondence with self-reported race/ethnicity**

Considering individual studies (Supplementary Table 2), SouthSeq and NYCKidSeq had the highest average African genetic ancestry, 32.8% (38.7%) and 28.7% (32.4%), respectively. KidsCanSeq and P^3^EGS had the highest average Native American genetic ancestry, 22.2% (25.2%) and 22.2% (27.7%) respectively, reflecting large numbers of Latino(a) cases (Table 1). European average genetic ancestry was high in all studies [min 35.1% (26.2%) in NYCKidSeq, max 82.5% (29.4%) in NCGENES 2]. Average East Asian genetic ancestry was < 10% in each study, but highest for P^3^EGS at 9.4% (25.5%). South Asian genetic ancestry was also < 10% in all studies, but highest in P^3^EGS and NYCKidSeq at 6.8% (20.4%) and 4.9% (SD=16.9%), respectively. Similarly, Middle Eastern genetic ancestry was also highest in P^3^EGS and NYCKidSeq at 10.0% (14.5%) and 14.4% (SD=16.7%), respectively.

There was correspondence between parent-reported or imputed race/ethnicity and estimated genetic ancestry of the unrelated cases (Supplementary Table 3). Cases described as 'White’ had 86.3% (18.4%) average European genetic ancestry and 10.2% (17.3%) average Middle Eastern genetic ancestry. Those described as Middle Eastern/North African/Mediterranean had 58.9% (37.9%) average Middle Eastern genetic ancestry, along with 12.3% (14.4%) average European and 23.1% (29.3%) average South Asian genetic ancestry. Cases described as Asian had 53.5% (46.2%) average East Asian and 42.6% (44.0%) average South Asian genetic ancestry across all studies. Supplementary Figure 4 shows that most Asian cases had either very high East Asian or South Asian genetic ancestry, but not admixed, across the five studies. Cases described as Black or African American had 83.0% (12.4%) average African genetic ancestry and 12.1% (9.4%) average European genetic ancestry. Cases described as Hispanic/Latino(a) had 41.8% (26.3%) average Native American and 30.8% (14.2%) average European genetic ancestry, as well as 13.2% (16.4%) average African and 12.6% (6.7%) average Middle Eastern genetic ancestry. Widespread Middle Eastern/Mediterranean genetic ancestry in Latino(a) individuals has been described previously^14,15^. Notably, the average Native-American and African genetic ancestries among reported Hispanic/Latino(a) cases varied by CSER study and were inversely correlated; for example, in P^3^EGS and KidsCanSeq the cases had the highest Native-American average genetic ancestry at 53.6% (4.9%) and 48.5% (16.6%) respectively, and the lowest average African genetic ancestry at 4.9% (6.1%) and 4.5% (4.7%) respectively, while in NYCKidSeq they had the lowest average Native-American genetic ancestry at 29.5% (30.2%) and highest average African genetic ancestry at 24.1% (19.3%) (Supplementary Tables 4, 6, 7), likely due to differences in nationalities. Cases described as having more than one race/ethnicity had high levels of genetic admixture (Supplementary Tables 3-8). Of note, those self-described as ‘American Indian/Native American/Alaska Native’ had 62.0% (39.5%) average European and 16.4% (29.3%) average Native American genetic ancestry (Supplementary Table 3).

#### **Genetic relatedness analysis findings**

Results from the relatedness analysis revealed seven genetically inferred full sibling pairs, two second-degree relative pairs (presumably half-sibs) and eleven third-degree relative pairs (presumed to be first cousins (Supplementary Figure 5). As expected, each pair of siblings shared the same study (two pairs from NYCKidSeq, SouthSeq, KidsCanSeq each and one from P^3^EGS). The two pairs of second-degree relatives were both from NYCKidSeq, while there were three pairs of third-degree relatives from NYCKidSeq, SouthSeq, P^3^EGS each. The remaining two pairs of third-degree relatives were from discordant studies, with one case from KidsCanSeq and the other from P^3^EGS each. All sibling pairs had similar case diagnoses and ancestry proportions. An unrelated set of cases was made by removing one member of each of the seven sibling pairs from the 3,015 cases for diagnostic yield analyses.

#### **Consanguinity, genetic ancestry, and diagnostic yield**

Overall, 14.6% of the 3008 unrelated cases had an F>= 0.0156, the expected level for offspring of second cousins (Supplementary Figure 7). Parental relatedness (consanguinity) increases the risk for AR inheritance^16–18^. As expected, there was a statistically significant increase in mean consanguinity in diagnosed AR homozygous cases (N=18) compared to undiagnosed cases in P^3^EGS (difference=0.044, *P*= 0.0004), while in NYCKidSeq and SouthSeq, there was also a trending but non-significant increase in mean consanguinity of diagnosed AR homozygous cases (N=6 and N=11 respectively) compared to undiagnosed cases. There were no statistical differences in consanguinity for any of the other modes of inheritance (Supplementary Tables 11-15).

Because diagnostic yield of ES in Middle Eastern and South Asian populations has been associated with consanguinity and AR inheritance^16–18^, we considered whether consanguinity in these ancestries (or others) impacted our diagnostic yield. We examined positive AR homozygous cases by consanguinity and genetic ancestry. From KidsCanSeq, NYCKidSeq, P^3^EGS and SouthSeq (the four studies with AR homozygous cases), 20/36 (54.3%) AR homozygous diagnosed cases had a consanguinity coefficient greater than 0.0156 (1/1 in KidsCanSeq, 14/18 in P^3^EGS, 3/6 in NYCKidSeq and 2/11 in SouthSeq). For these 20 cases, mean (SD) genetic ancestries were: 7.3% (20.1%) African ancestry, 23.4% (34.6%) Native American ancestry, 5.6% (19.6%) East Asian ancestry, 18.2% (23.5%) European ancestry, 15.1% (29.0%) Middle Eastern Ancestry, 30.0% (41.1%) South Asian Ancestry. For the 16 cases with F<0.0156, mean genetic ancestries were 30.8% (35.6%) African ancestry, 17.5% (34.8%) Native American ancestry, 1.4% (2.8%) East Asian ancestry, 39.4% (35.9%) European ancestry, 6.1% (7.6%) Middle Eastern Ancestry, 4.7% (12.7%) South Asian Ancestry. The increase of South Asian and Middle Eastern ancestries among those with consanguinity is evident, consistent with prior studies^16–18^. However, ancestry-related homozygous inheritance due to consanguinity did not impact the overall diagnostic yield.

#### **Pathogenic/Likely Pathogenic (P/LP) variant types, and variant allele frequencies in positive cases**

Frameshift, missense, splice-site, stop-gain P/LP variant types differed in frequency among the KidsCanSeq, NYCKidSeq, P^3^EGS, and SouthSeq studies (Chi-square statistic: 19.15, *P=*0.024; Supplementary Table 16). This association was largely driven by differences in splice-site variant proportions across studies, with lower frequencies in NYCKidSeq and P^3^EGS and higher frequencies in KidsCanSeq and SouthSeq. Frameshift variants were most common in KidsCanSeq, while missense variants were most common for the other three studies. The rest of the variant types (start lost, in-frame deletion, in-frame insertion, deletion-insertion/insertion-deletion and CNVs) were infrequent except for CNVs in NYCKidSeq and SouthSeq. Both had high proportions of P/LP variants that were CNVs (27.6%, 29.3% respectively) due to their use of genome sequencing and no prior array analyses, while no other study had >1% proportion of CNVs.

P/LP variants in positive cases had mostly very low allele frequencies (567/575 P/LP variants had an ancestry specific allele frequency in gnomAD V4 <0.001) or were missing from gnomAD V4 database altogether (Supplementary Table 17).

#### **Recurrent variants across studies**

We found five recurrent variants (each occurring twice) in three different genes among 10 different positive cases; three different recurrent variants were in *PTPN11* (Supplementary Table 18). All the recurrent variants were AD; seven were *de novo* (all found in P^3^EGS), one was inheritance unknown (in P^3^EGS), and the remaining two were inherited (maternal in SouthSeq and paternal in NYCKidSeq). No recurrent inherited founder variants (either AR or AD) were observed.

### **Supplementary Figures**

#### **Supplementary Figure 1: First and second genetic ancestry principal components (PC 1 and PC 2) of CSER participants with HGDP samples projected**

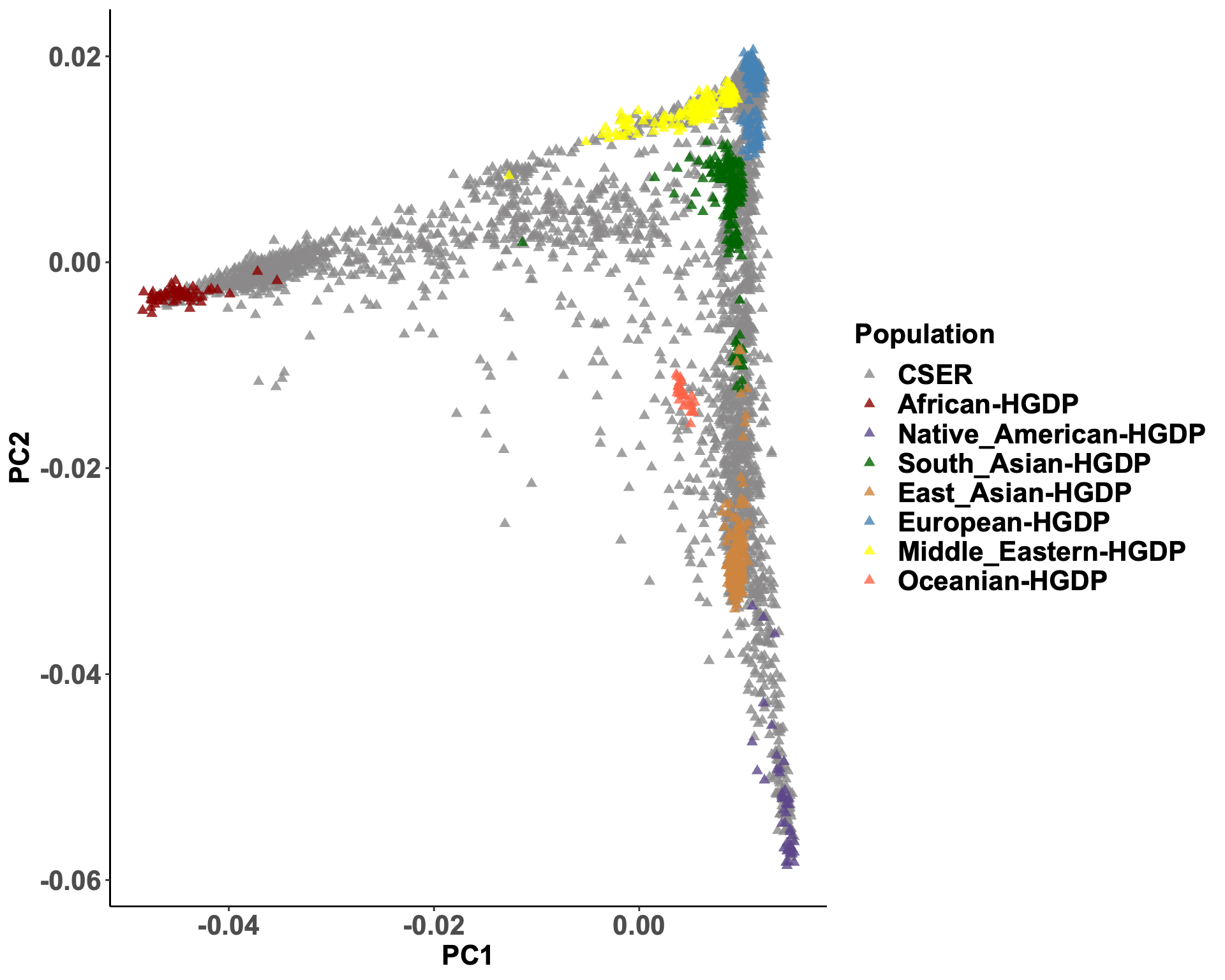

#### **Supplementary Figure 2: Third and fourth genetic ancestry principal components (PC 3 and PC 4) of CSER participants with HGDP samples projected**

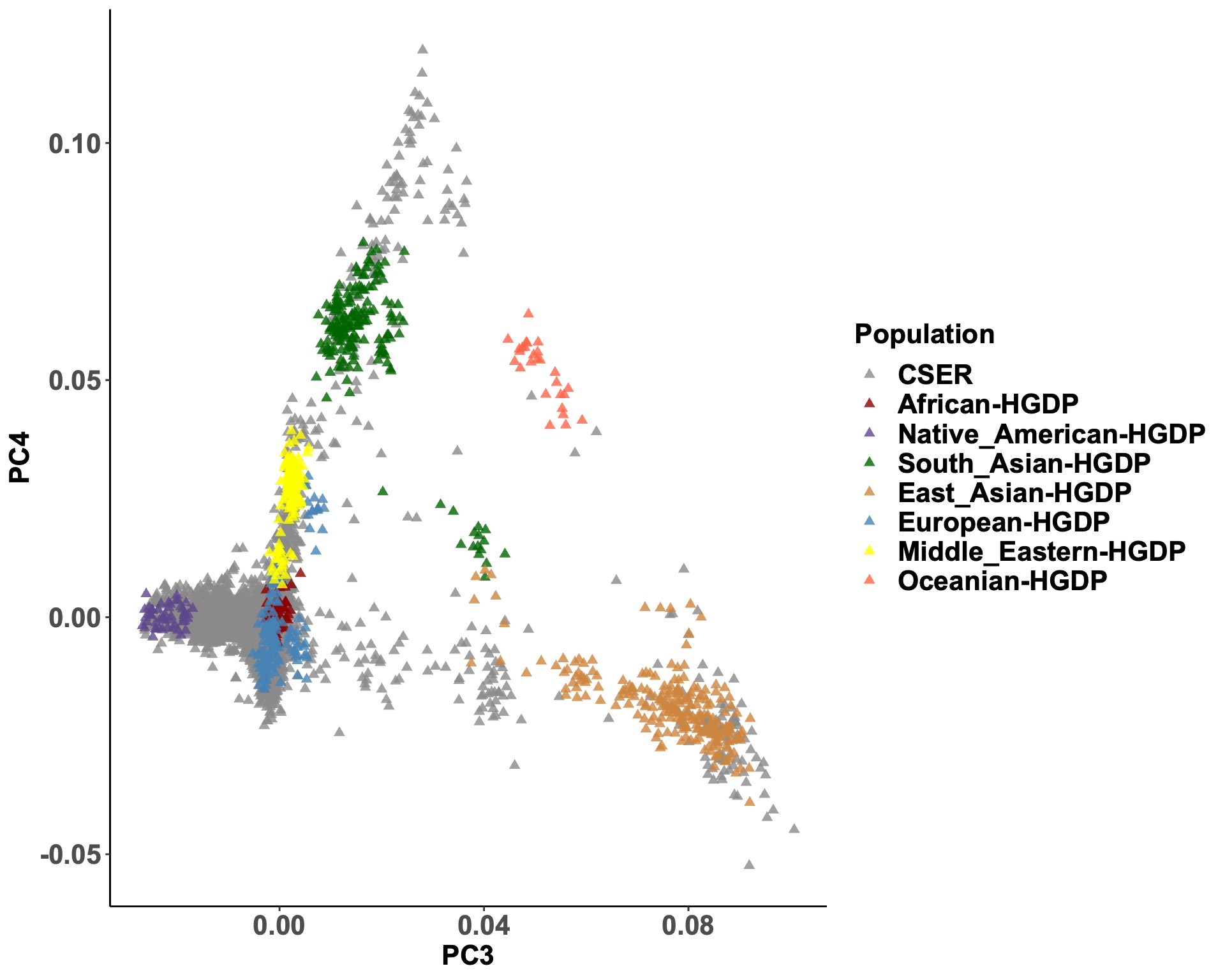

#### **Supplementary Figure 3: Fifth and sixth genetic ancestry principal components (PC 5 and PC 6) of CSER participants with HGDP samples projected**

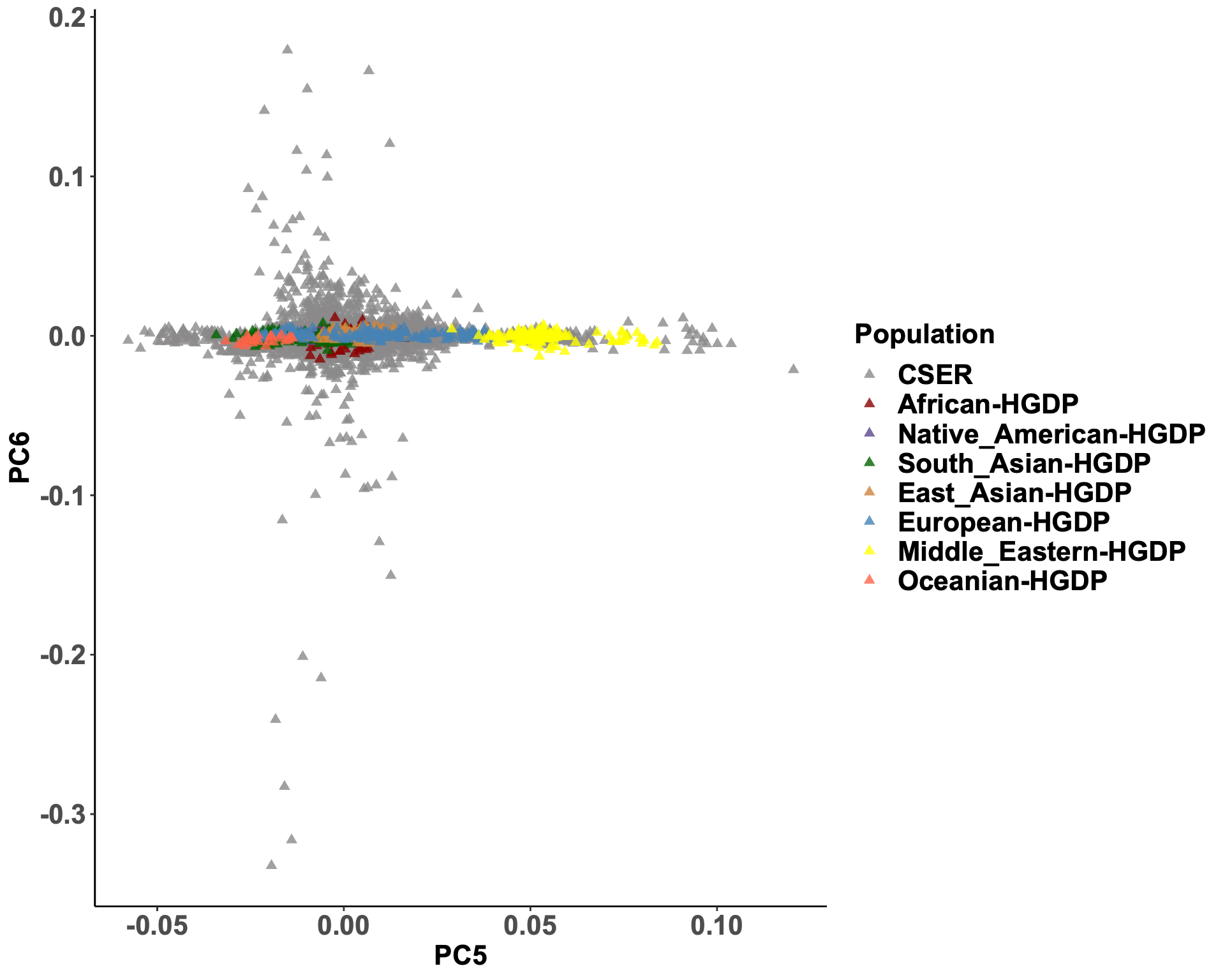

#### **Supplementary Figure 4: Estimated Global Genetic Ancestry/Admixture of Cases in the 5 CSER Sites.**

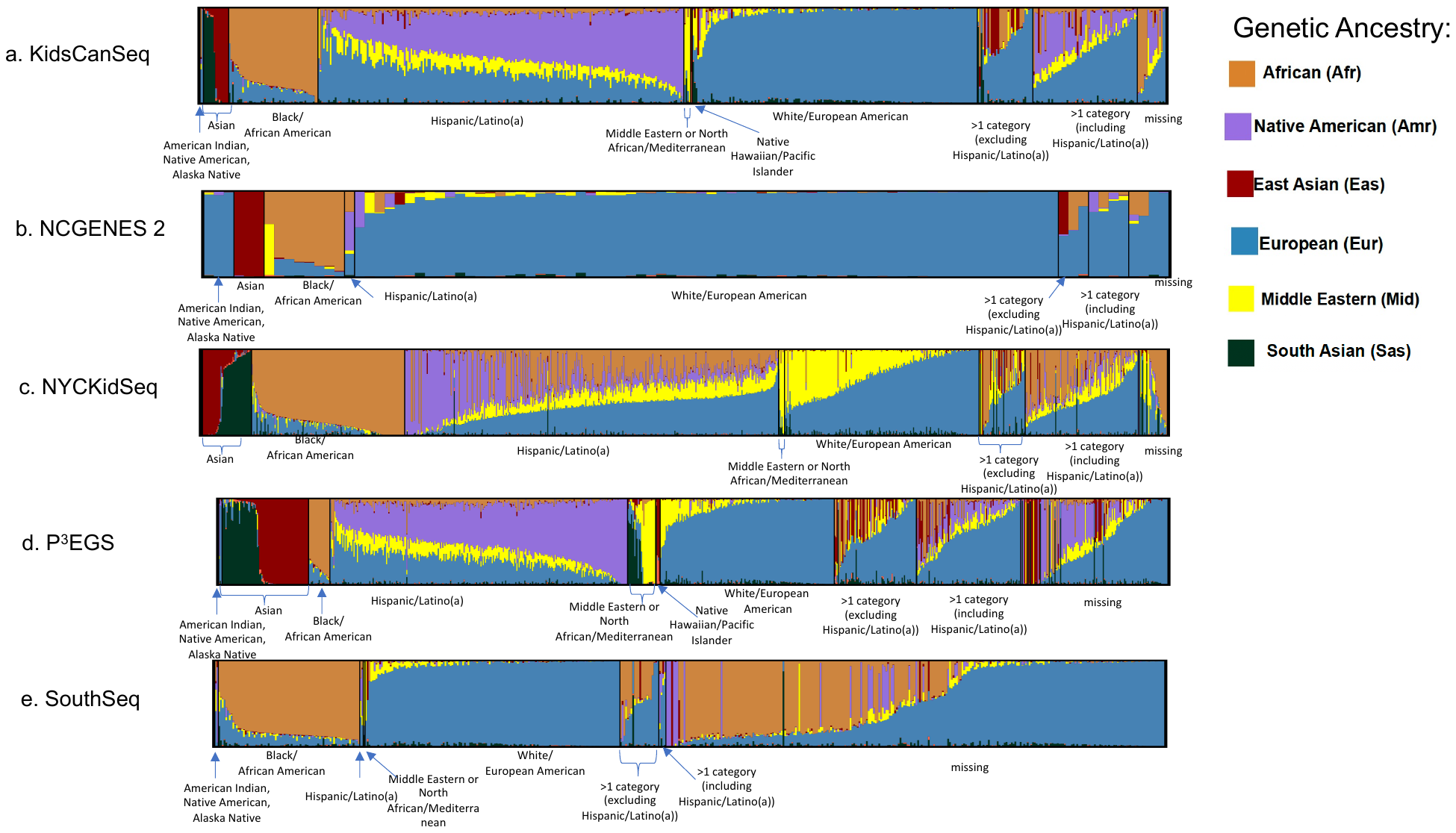

Estimated global genetic ancestry proportions for cases are shown for each CSER site. Each vertical bar on the x-axis represents an individual case, while the y-axis indicates the proportion of genetic ancestry, ranging from 0% to 100%. The colors within each bar represent the percentage of genetic ancestry attributed to different groups (African, Native American, East Asian, European, Middle Eastern, Oceanian, South Asian).

These proportions were calculated using sequencing data analyzed with the ADMIXTURE software, referencing unrelated Human Genome Diversity Panel (HGDP) samples from gnomAD. Panels (a-e) correspond to cases from KidsCanSeq, NCGENES 2, NYCKidSeq, P3EGS, and SouthSeq, respectively. The height and color distribution within each bar reflect the genetic ancestry composition of individuals in each site.

**Supplementary Figure 5: Identification of siblings/ sibling pairs across CSER studies using PCRelate.**

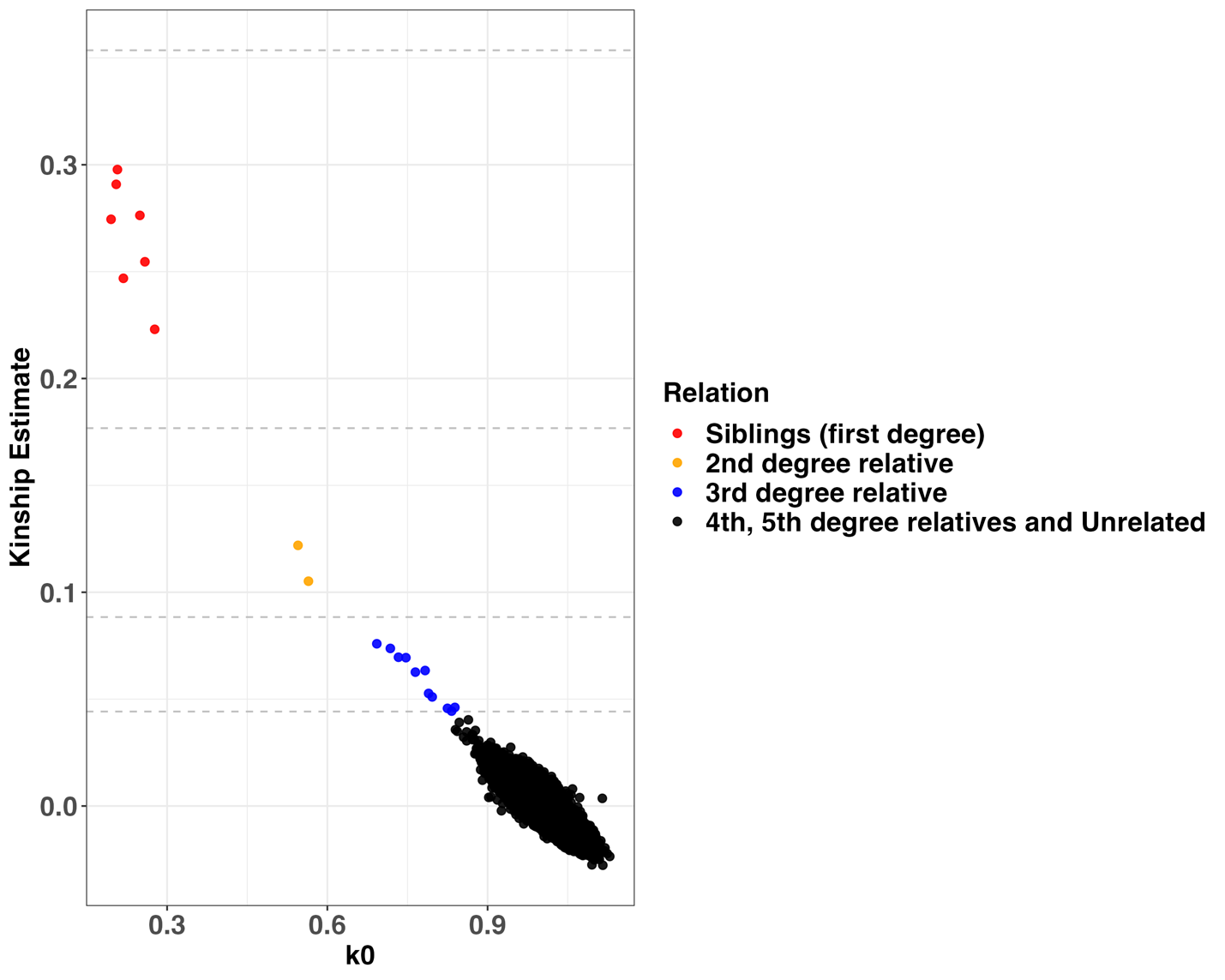

Each dot represents a pair of individuals among the 3015 sequenced cases. The Y axis shows the kinship coefficient, which measures how genetically similar two individuals are. Higher values mean the individuals are more closely related. The x-axis (k0) shows the chance that any shared allele between two individuals is identical by state (IBS). IBS means the alleles are the same, whether they came from a common ancestor or not.

#### **Supplementary Figure 6: Distribution of Consanguinity Coefficients by CSER Study**

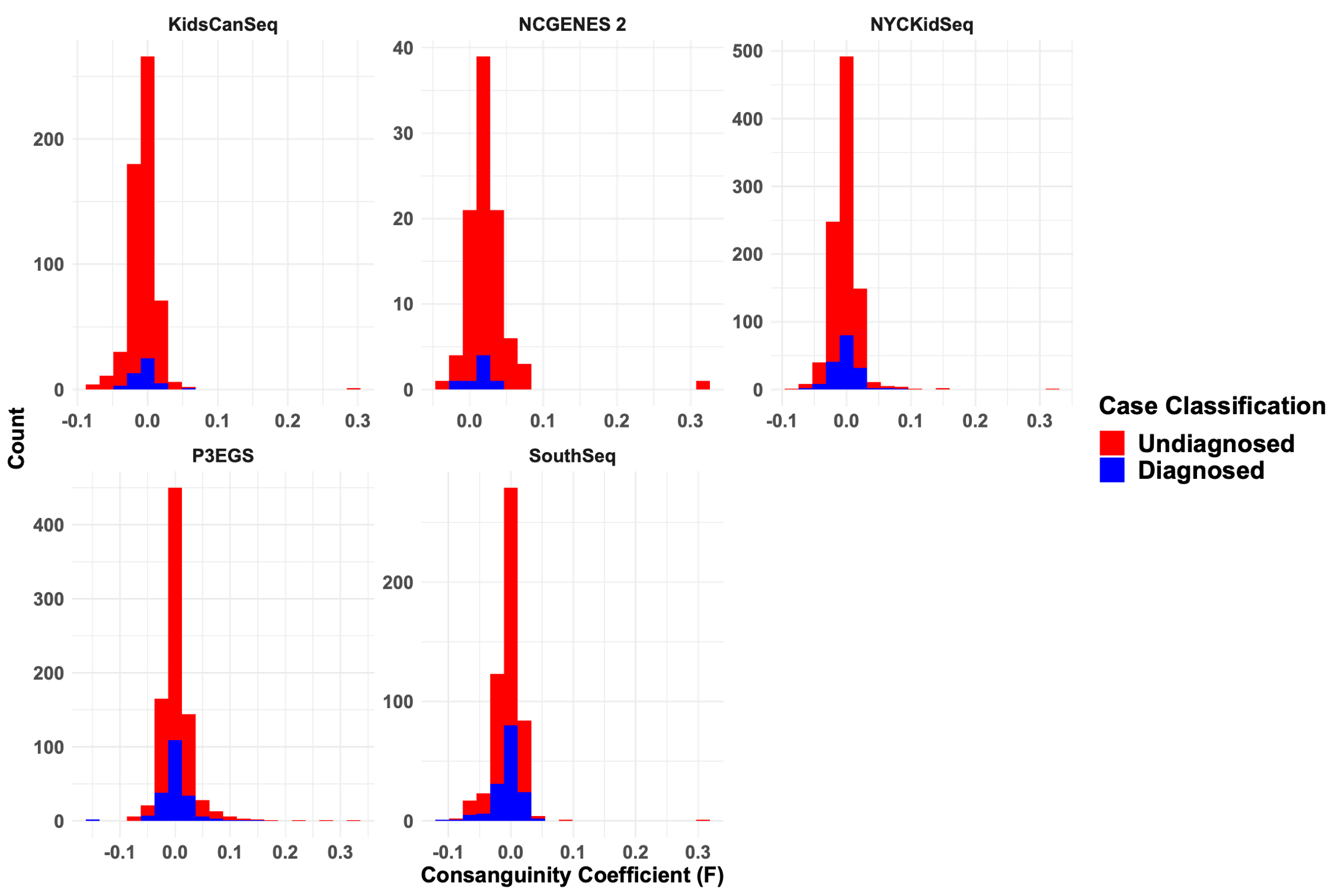

Distribution of consanguinity coefficients ($F$) across CSER studies. Red histograms represent undiagnosed cases and blue histograms represent diagnosed cases.

**Supplementary Figure 7: Association of Study Factors with Diagnostic Yield in Autosomal Dominant Mode of Inheritance**
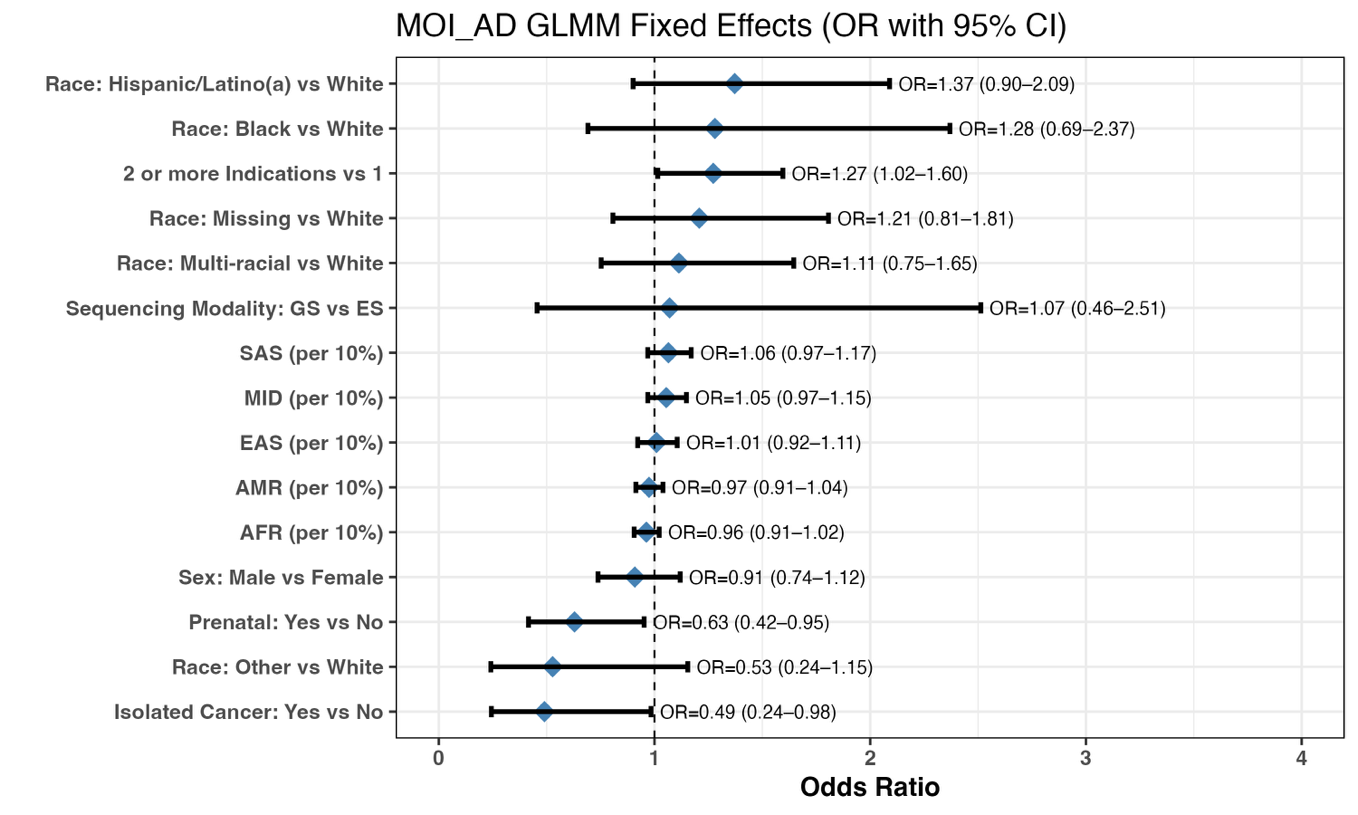

Forest plot showing odds ratios (ORs) and 95% confidence intervals for fixed effects from the generalized linear mixed model in Autosomal Dominant cases vs Undiagnosed. Covariates include genetic ancestry (per 10% increase), demographic and clinical characteristics, and sequencing modality, with adjustment for study site as a random effect. Odds ratios greater than 1 indicate higher odds of a positive diagnostic yield.

#### **Supplementary Figure 8: Association of Study Factors with Diagnostic Yield in Autosomal Recessive Mode of Inheritance**

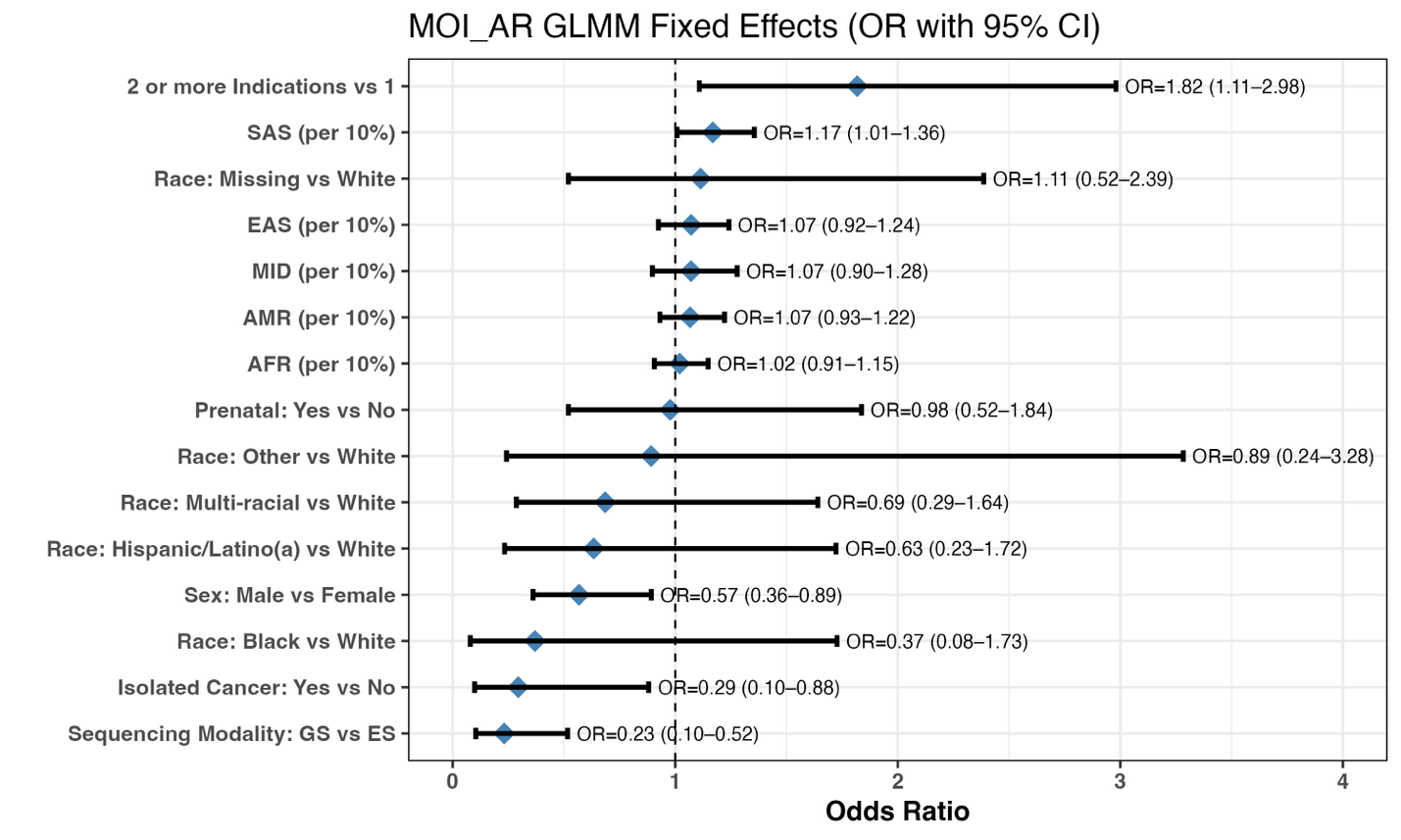

Forest plot showing odds ratios (ORs) and 95% confidence intervals for fixed effects from the generalized linear mixed model in Autosomal Recessive cases vs Undiagnosed. Covariates include genetic ancestry (per 10% increase), demographic and clinical characteristics, and sequencing modality, with adjustment for study site as a random effect. Odds ratios greater than 1 indicate higher odds of a positive diagnostic yield.

#### **Supplementary Figure 9: Association of Study Factors with Diagnostic Yield in X-Linked Mode of Inheritance**

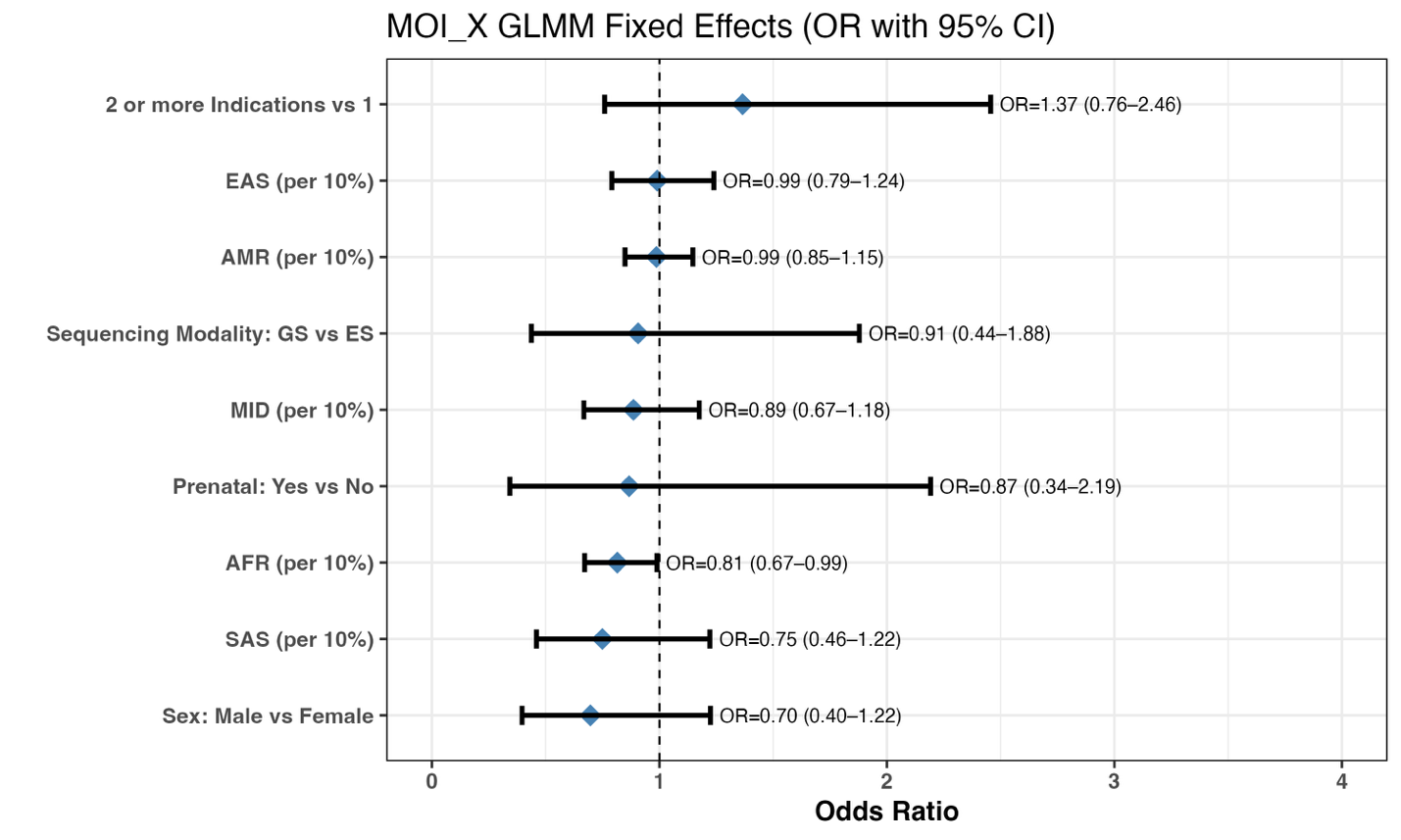

Forest plot showing odds ratios (ORs) and 95% confidence intervals for fixed effects from the generalized linear mixed model in X-Linked cases vs Undiagnosed. Covariates include genetic ancestry (per 10% increase), demographic and clinical characteristics, and sequencing modality, with adjustment for study site as a random effect. Odds ratios greater than 1 indicate higher odds of a positive diagnostic yield.

### **Supplementary Tables**

#### **Supplementary Table 1: Self-identified race/ethnicity of parents of cases in the 5 centers**

|  | **NCGENES 2** | **P^3^EGS** | | | **KidsCanSeq** | **NYCKidSeq** | | | **SouthSeq** | **Total** |
| --- | --- | --- | --- | --- | --- | --- | --- | --- | --- | --- |
| **Number of Parents responding** | 1 | 2 | | | 1 | 2 | | | 1 |  |
| **Parent a, b or average frequency** | Parent a | Parent a | Parent b | Average | Parent a | Parent a | Parent b | Average | Parent a |  |
| **Race/Ethnicity N (column %)** |  |  |  |  |  |  |  |  |  |  |
| American Indian, Native American, Alaska Native (NAT) | 3  (3.1) | 5  (0.6) | 5  (0.6) | 5  (0.6) | 3  (0.5) | 4  (0.4) | 2  (0.2) | 3  (0.3) | 4  (0.7) | 18  (0.6) |
| Asian (AS) | 0  (0.0) | 92  (10.9) | 81  (9.6) | 86.5  (10.3) | 20  (3.5) | 54  (5.6) | 52  (5.4) | 53  (5.5) | 3  (0.6) | 162.5 (5.4) |
| Black or African American (AF) | 8  (8.3) | 24  (2.8) | 29  (3.4) | 26.5  (3.1) | 52  (9.1) | 153 (15.9) | 165  (17.1) | 159 (16.5) | 80  (14.9) | 325.5 (10.8) |
| Native Hawaiian/Pacific Islander (PI) | 0  (0.0) | 5  (0.6) | 6  (0.7) | 5.5  (0.7) | 1  (0.2) | 0  (0) | 0  (0) | 0  (0) | 0  (0) | 6.5  (0.2) |
| White or European American (EU) | 77  (80.2) | 219  (25.9) | 215  (25.4) | 217  (25.7) | 202  (35.3) | 216 (22.4) | 220  (22.8) | 218 (22.6) | 154 (28.7) | 868 (28.8) |
| Middle Eastern or North African/Mediterranean (ME) | 0  (0.0) | 28  (3.3) | 29  (3.4) | 28.5  (3.5) | 5  (0.9) | 12  (1.2) | 11  (1.1) | 11.5 (1.2) | 1  (0.2) | 46  (1.5) |
| Hispanic/Latino(a) (LT) | 1  (1.0) | 282  (33.4) | 260  (30.8) | 271  (32.1) | 221  (38.6) | 390 (40.5) | 379  (39.3) | 384.5 (39.9) | 4  (0.7) | 881.5 (29.2) |
| Prefer not to answer | 0  (0.0) | 6  (0.7) | 7  (0.8) | 6.5  (0.8) | 8  (1.4) | 12  (1.2) | 12  (1.2) | 12  (1.2) | 1  (0.2) | 27.5 (0.9) |
| Unknown/none of these fully describe me | 1  (1.0) | 2  (0.2) | 0  (0.0) | 1  (0.1) | 5  (0.9) | 14  (1.5) | 25  (2.6) | 19.5 (2.0) | 0  (0.0) | 26.5 (0.9) |
| Not Reported | 2  (2.1) | 113  (13.4) | 166  (19.6) | 139.5  (16.5) | 10  (1.7) | 44  (4.6) | 44  (4.6) | 44  (4.6) | 277 (51.6) | 472.5 (15.7) |
| AF, EU | 1  (1.0) | 6  (0.7) | 2  (0.2) | 4  (0.5) | 1  (0.2) | 1  (0.1) | 2  (0.2) | 1.5  (0.2) | 6  (1.1) | 13.5 (0.5) |
| AF, LT | 0  (0.0) | 4  (0.5) | 2  (0.2) | 3  (0.4) | 3  (0.5) | 16  (1.7) | 16  (1.7) | 16  (1.7) | 0  (0.0) | 22  (0.7) |
| AS, EU | 0  (0.0) | 10  (1.2) | 2  (0.2) | 6  (0.7) | 5  (0.9) | 5  (0.5) | 2  (0.2) | 3.5  (0.4) | 0  (0.0) | 14.5 (0.5) |
| EU, LT | 3  (3.1) | 15  (1.8) | 20  (2.4) | 17.5  (2.1) | 26  (4.5) | 22  (2.3) | 14  (1.5) | 18  (1.9) | 2  (0.4) | 66.5 (2.2) |
| Other combinations | 0  (0.0) | 34  (4.0) | 21  (2.5) | 27.5  (3.3) | 11  (1.9) | 21  (2.2) | 20  (2.1) | 20.5 (2.1) | 5  (0.9) | 64  (2.1) |
| Total | 96 | 845 | 845 | 845 | 573 | 964 | 964 | 964 | 537 | 3015 |

For NCGENES 2, KidsCanSeq and SouthSeq, only one parent reported their R/E, while for P^3^EGS and NYCKidSeq both parents reported. To calculate the average and then the total, a weight of 1/2 was applied for each of the parents where there are two parents (P^3^EGS and NYCKidSeq)

#### **Supplementary Table 2: Mean and standard deviation (SD) of estimated genetic ancestries by center and overall**

| **Genetic Ancestry** | **KidsCanSeq**  **(N=573)** | **NCGENES 2**  **(N=96)** | **NYCKidSeq**  **(N=964)** | **P^3^EGS**  **(N=845)** | **SouthSeq**  **(N=537)** | **Overall**  **(N=3015)** |
| --- | --- | --- | --- | --- | --- | --- |
| African | 0.120  (0.247) | 0.088  (0.227) | 0.287  (0.324) | 0.063  (0.164) | 0.328  (0.387) | 0.194  (0.305) |
| Native American | 0.222  (0.252) | 0.019  (0.069) | 0.132  (0.237) | 0.222  (0.277) | 0.041  (0.155) | 0.155  (0.247) |
| East Asian | 0.031  (0.134) | 0.039  (0.180) | 0.032  (0.144) | 0.094  (0.255) | 0.010  (0.061) | 0.045  (0.176) |
| European | 0.525  (0.326) | 0.825  (0.294) | 0.351  (0.262) | 0.448  (0.332) | 0.574  (0.392) | 0.466  (0.338) |
| Middle Eastern | 0.072  (0.098) | 0.020  (0.069) | 0.144  (0.167) | 0.100  (0.145) | 0.027  (0.067) | 0.093  (0.139) |
| South Asian | 0.026  (0.110) | 0.006  (0.012) | 0.049  (0.169) | 0.068  (0.204) | 0.017  (0.050) | 0.043  (0.155) |

#### **Supplementary Table 3: Mean and standard deviation (SD) of estimated genetic ancestry% by race/ethnicity in unrelated cases across all centers**

| **Cases’ race/ethnicity** | **Count** | **African** | **Native American** | **East Asian** | **European** | **Middle Eastern** | **South Asian** | **Oceanian** |
| --- | --- | --- | --- | --- | --- | --- | --- | --- |
| American Indian, Native American, Alaska Native | **11** | 7.08 (16.50) | 16.40 (29.30) | 3.68  (5.80) | 62.00  (39.50) | 2.84 (6.29) | 7.58 (23.30) | 0.43 (0.73) |
| Asian | **143** | 0.01 (0.08) | 0.28 (0.85) | 53.30 (46.20) | 2.29 (6.95) | 0.39 (2.06) | 42.60  (44.00) | 1.05 (1.44) |
| Black or African American | **313** | 83.00 (12.40) | 0.71 (1.85) | 0.40  (0.88) | 12.10 (9.36) | 1.87 (4.85) | 1.55 (3.28) | 0.36 (0.54) |
| Hispanic/Latino(a) | **854** | 13.20 (16.40) | 41.80 (26.30) | 0.46 (1.44) | 30.80 (14.20) | 12.60 (6.66) | 0.84 (2.40) | 0.33 (0.53) |
| Middle Eastern or North African/Mediterranean | **36** | 2.99 (6.41) | 0.28 (0.98) | 1.94 (4.54) | 12.30 (14.40) | 58.90 (37.90) | 23.10 (29.30) | 0.40  (0.66) |
| Native Hawaiian/Pacific Islander | **5** | 0.00  (0.00) | 0.00  (0.00) | 60.30 (34.30) | 0.68 (1.52) | 0.00  (0.00) | 18.50 (41.30) | 20.60 (15.10) |
| White or European American | **732** | 0.41 (2.46) | 0.70  (3.13) | 0.42 (2.77) | 86.30 (18.40) | 10.20 (17.30) | 1.76 (2.94) | 0.25 (0.43) |
| More than one category (excluding Hispanic/Latino(a)) | **177** | 18.00 (25.50) | 1.91 (7.00) | 11.60 (19.70) | 52.80 (24.70) | 9.07 (15.60) | 5.83 (14.80) | 0.74 (1.74) |
| More than one category (including Hispanic/Latino(a)) | **276** | 17.60 (22.90) | 16.50 (15.10) | 3.87 (11.40) | 49.80 (22.50) | 9.93 (8.73) | 1.77 (4.86) | 0.44 (0.64) |
| Not Reported | **408** | 26.40 (36.50) | 11.20 (24.20) | 3.96 (16.50) | 50.30 (37.70) | 4.40 (9.52) | 3.36 (12.90) | 0.35  (0.60) |
| Prefer not to answer | **27** | 37.00 (39.10) | 12.00  (22.70) | 1.29 (3.17) | 30.20 (26.50) | 17.30 (25.30) | 1.78 (5.82) | 0.43 (0.51) |
| Unknown/none of these fully describe me | **26** | 24.30 (23.60) | 4.34 (9.93) | 1.25 (2.96) | 42.90 (27.10) | 12.40  (23.00) | 14.30 (30.10) | 0.54 (0.85) |
| Overall | **3008** | 19.30 (30.50) | 15.50  (24.70) | 4.54  (17.60) | 46.60  (33.80) | 9.30  (13.90) | 4.29 (15.40) | 0.42  (1.24) |

#### **Supplementary Table 4: Mean and standard deviation (SD) of estimated genetic ancestry % by race/ethnicity in unrelated KidsCanSeq cases**

| **Cases’ race/ethnicity** | **Count** | **African** | **Native American** | **East Asian** | **European** | **Middle Eastern** | **South Asian** | **Oceanian** |
| --- | --- | --- | --- | --- | --- | --- | --- | --- |
| American Indian, Native American, Alaska Native | **2** | 25.90 (36.60) | 3.46  (2.75) | 1.41 (0.74) | 66.90  (43.30) | 2.27  (3.21) | 0.00  (0.00) | 0.01  (0.01) |
| Asian | **15** | 0.00  (0.00) | 0.06  (0.19) | 56.30 (46.30) | 0.99  (2.67) | 0.00  (0.00) | 41.40 (45.90) | 1.25  (1.31) |
| Black or African American | **53** | 79.50 (11.30) | 0.73  (1.54) | 0.62 (1.51) | 16.70  (9.75) | 0.59  (1.16) | 1.51 (3.26) | 0.35  (0.54) |
| Hispanic/Latino(a) | **215** | 4.48 (4.71) | 48.50  (16.60) | 0.52 (1.79) | 33.90  (13.60) | 11.60  (6.30) | 0.76 (1.37) | 0.33  (0.51) |
| Middle Eastern or North African/Mediterranean | **4** | 6.09 (7.65) | 0.00  (0.00) | 0.21 (0.41) | 10.20  (8.35) | 76.80  (16.80) | 6.27 (12.40) | 0.47  (0.33) |
| More than one category (excluding Hispanic/Latino(a)) | **33** | 13.30 (18.90) | 1.72  (6.56) | 13.60 (19.30) | 60.30  (23.30) | 3.86  (7.65) | 6.69 (18.40) | 0.59  (0.86) |
| More than one category (including Hispanic/Latino(a)) | **62** | 8.35 (14.30) | 27.40  (17.70) | 2.43 (7.63) | 52.20  (21.30) | 8.16  (6.45) | 1.12 (1.66) | 0.42  (0.63) |
| Native Hawaiian/Pacific Islander | **1** | 0.00  (NA) | 0.00  (NA) | 67.3 (NA) | 0.00  (NA) | 0.00  (NA) | 0.00  (NA) | 32.7  (NA) |
| Not Reported | **4** | 26.40 (38.70) | 21.70  (25.60) | 0.29 (0.58) | 45.70  (37.00) | 5.81  (5.00) | 0.00  (0.00) | 0.12  (0.24) |
| Prefer not to answer | **9** | 46.40 (41.40) | 17.20  (19.90) | 1.72 (4.27) | 23.70  (12.60) | 9.60  (13.50) | 0.99 (1.20) | 0.42  (0.45) |
| Unknown/none of these fully describe me | **4** | 23.50 (26.60) | 7.79  (13.40) | 0.21 (0.22) | 59.90  (23.80) | 7.44  (11.30) | 0.87 (0.57) | 0.25  (0.40) |
| White or European American | **169** | 0.29 (1.11) | 0.88  (3.24) | 0.59 (3.92) | 93.30  (11.90) | 2.89  (7.31) | 1.76 (4.11) | 0.24  (0.43) |
| Overall | **571** | 12.00 (24.80) | 22.10  (25.30) | 3.11 (13.40) | 52.50  (32.60) | 7.16  (9.82) | 2.61 (11.00) | 0.52  (0.41) |

#### **Supplementary Table 5: Mean and standard deviation (SD) of estimated genetic ancestry % by race/ethnicity in unrelated NCGENES 2 cases**

| **Cases’ race/ethnicity** | **Count** | **African** | **Native American** | **East Asian** | **European** | **Middle Eastern** | **South Asian** | **Oceanian** |
| --- | --- | --- | --- | --- | --- | --- | --- | --- |
| American Indian, Native American, Alaska Native | **3** | 0.29 (0.51) | 1.44  (2.27) | 0.19 (0.32) | 96.60  (2.53) | 0.80  (1.38) | 0.49 (0.85) | 0.18  (0.22) |
| Asian | **3** | 0.00  (0.00) | 0.50  (0.86) | 99.10 (0.49) | 0.00  (0.00) | 0.00  (0.00) | 0.00  (0.00) | 0.40  (0.40) |
| Black or African American | **8** | 78.20 (17.00) | 0.19  (0.36) | 0.59 (0.52) | 12.20  (7.00) | 7.76  (21.30) | 0.39 (0.57) | 0.68  (0.63) |
| Hispanic/Latino(a) | **1** | 23.70 (NA) | 45.60  (NA) | 0.00  (NA) | 24.10  (NA) | 4.38  (NA) | 2.06 (NA) | 0.16  (NA) |
| More than one category (excluding Hispanic/Latino(a)) | **3** | 20.70 (22.70) | 0.78  (0.77) | 16.60 (28.80) | 61.00  (16.90) | 0.00  (0.00) | 0.35 (0.60) | 0.51  (0.88) |
| More than one category (including Hispanic/Latino(a)) | **4** | 5.79 (8.72) | 8.44  (10.90) | 0.13 (0.26) | 83.60  (7.65) | 1.67  (2.18) | 0.00  (0.00) | 0.32  (0.64) |
| Not Reported | **2** | 0.00  (0.00) | 0.53  (0.74) | 0.05 (0.07) | 98.20  (0.92) | 0.00  (0.00) | 1.23 (1.74) | 0.00  (0.00) |
| Unknown/none of these fully describe me | **2** | 27.70 (1.02) | 3.49  (4.94) | 0.00  (0.00) | 67.40  (5.39) | 1.10  (1.55) | 0.02 (0.02) | 0.35  (0.05) |
| White or European American | **70** | 0.74 (3.00) | 1.14  (5.35) | 0.36 (1.80) | 95.10  (7.00) | 1.65  (3.80) | 0.67 (1.29) | 0.33  (0.5) |
| Overall | **96** | 8.77 (22.70) | 1.85  (6.91) | 3.94 (1.80) | 82.50  (29.40) | 2.01  (6.90) | 0.59 (1.16) | 0.35  (0.50) |

#### **Supplementary Table 6: Mean and standard deviation (SD) of estimated genetic ancestry % by race/ethnicity in unrelated NYCKidSeq cases**

| **Cases’ race/ethnicity** | **Count** | **African** | **Native American** | **East Asian** | **European** | **Middle Eastern** | **South Asian** | **Oceanian** |
| --- | --- | --- | --- | --- | --- | --- | --- | --- |
| American Indian, Native American, Alaska Native | **1** | 0.00  (NA) | 0.28  (NA) | 19.70 (NA) | 0.00  (NA) | 0.00  (NA) | 77.80 (NA) | 2.22  (NA) |
| Asian | **49** | 0.00  (0.00) | 0.42  (1.10) | 41.50 (43.30) | 2.82  (9.50) | 0.28  (1.91) | 53.70 (42.40) | 1.22  (1.51) |
| Black or African American | **153** | 85.00  (13.00) | 0.85  (2.30) | 0.28 (0.59) | 9.93  (9.00) | 1.97  (4.25) | 1.65 (3.70) | 0.33  (0.55) |
| Hispanic/Latino(a) | **371** | 24.10 (19.30) | 29.50  (30.20) | 0.43 (1.52) | 31.30  (15.00) | 13.50  (7.13) | 0.85 (3.30) | 0.34  (0.54) |
| Middle Eastern or North African/Mediterranean | **6** | 3.14 (5.80) | 0.00  (0.00) | 0.13 (0.31) | 8.98  (10.50) | 74.10  (27.70) | 13.40 (20.20) | 0.27  (0.33) |
| More than one category (excluding Hispanic/Latino(a)) | **46** | 23.00  (30.10) | 1.03  (4.60) | 10.60 (21.00) | 40.10  (20.90) | 16.40  (23.30) | 8.22 (17.90) | 0.60  (0.91) |
| More than one category (including Hispanic/Latino(a)) | **113** | 30.90 (25.70) | 10.90  (11.60) | 2.19 (8.10) | 40.50  (19.00) | 13.30  (10.40) | 1.72 (6.00) | 0.44  (0.67) |
| Not Reported | **2** | 40.70 (57.60) | 50.00  (70.70) | 0.00  (0.00) | 7.82  (11.10) | 0.00  (0.00) | 1.15 (1.62) | 0.31  (0.43) |
| Prefer not to answer | **12** | 47.90 (38.10) | 5.18  (10.10) | 0.38 (0.75) | 23.50  (16.40) | 19.60  (24.20) | 2.89 (8.70) | 0.57  (0.59) |
| Unknown/none of these fully describe me | **15** | 22.30 (26.60) | 4.92  (11.20) | 1.79 (3.78) | 27.40  (21.60) | 18.80  (28.30) | 24.20 (32.10) | 0.66  (0.96) |
| White or European American | **194** | 0.58 (3.90) | 0.60  (3.60) | 0.37 (1.89) | 68.90  (23.00) | 27.20  (22.60) | 2.12 (3.10) | 0.19  (0.37) |
| Overall | **962** | 28.70 (32.40) | 13.20  (23.70) | 3.22 (14.40) | 35.20  (26.20) | 14.40  (16.70) | 4.93 (16.90) | 0.39  (0.68) |

#### **Supplementary Table 7: Mean and standard deviation (SD) of estimated genetic ancestry % by race/ethnicity in unrelated P^3^EGS cases**

| **Combined parents’ race/ethnicity** | **Count** | **African** | **Native American** | **East Asian** | **European** | **Middle Eastern** | **South Asian** | **Oceanian** |
| --- | --- | --- | --- | --- | --- | --- | --- | --- |
| American Indian, Native American, Alaska Native | **3** | 0.42 (0.37) | 24.90  (31.30) | 2.51 (2.71) | 69.40  (35.50) | 1.06  (1.83) | 1.10  (1.90) | 0.65  (0.7) |
| Asian | **76** | 0.01 (0.11) | 0.22  (0.74) | 58.50 (47.30) | 2.30  (5.61) | 0.56  (2.37) | 37.40 (44.10) | 0.92  (1.50) |
| Black or African American | **19** | 81.70 (7.24) | 0.39  (0.71) | 0.59 (1.00) | 14.00  (7.36) | 1.77  (2.04) | 0.92 (1.36) | 0.68  (0.48) |
| Hispanic/Latino(a) | **265** | 4.90  (6.10) | 53.60  (18.50) | 0.47 (0.93) | 27.60  (13.10) | 12.20  (6.10) | 0.90  (1.43) | 0.31  (0.53) |
| Middle Eastern or North African/Mediterranean | **25** | 2.56 (6.60) | 0.41  (1.16) | 2.73 (5.28) | 13.60  (16.30) | 51.80  (41.50) | 28.60 (32.3) | 0.43  (0.76) |
| More than one category (excluding Hispanic/Latino(a)) | **73** | 10.40 (20.50) | 2.30  (7.86) | 13.90 (20.50) | 58.40  (24.40) | 9.24  (12.70) | 4.75 (11.30) | 0.96  (2.50) |
| More than one category (including Hispanic/Latino(a)) | **93** | 8.68 (16.20) | 16.10  (12.60) | 7.00 (16.00) | 57.90  (22.80) | 7.50  (6.46) | 2.28 (4.94) | 0.47  (0.63) |
| Native Hawaiian/Pacific Islander | **4** | 0.00  (0.00) | 0.00  (0.00) | 58.50 (39.40) | 0.85  (1.70) | 0.00  (0.00) | 23.10 (46.20) | 17.50  (15.60) |
| Not Reported | **125** | 6.29 (18.00) | 20.90  (28.70) | 10.40 (26.60) | 46.20  (34.20) | 8.67  (13.70) | 7.12 (21.30) | 0.48  (0.76) |
| Prefer not to answer | **5** | 1.06 (1.46) | 21.30  (43.90) | 2.98 (4.54) | 44.50  (42.80) | 29.20  (42.40) | 0.71 (21.30) | 0.19  (0.42) |
| Unknown/none of these fully describe me | **1** | 0.00  (NA) | 0.00  (NA) | 0.00  (NA) | 97.50  (NA) | 0.00  (NA) | 2.50  (NA) | 0.00  (NA) |
| White or European American | **155** | 0.19 (0.77) | 0.70  (2.10) | 0.24 (0.92) | 88.90  (12.90) | 7.66  (12.40) | 2.07 (2.51) | 0.25  (0.44) |
| Overall | **844** | 6.29 (16.40) | 22.30  (27.70) | 9.37 (25.50) | 44.80  (33.20) | 9.96  (14.60) | 6.74 (20.30) | 0.54  (1.82) |

#### **Supplementary Table 8: Mean and standard deviation (SD) of estimated genetic ancestry % by race/ethnicity in unrelated SouthSeq cases**

| **Cases’ race/ethnicity** | **Count** | **African** | **Native American** | **East Asian** | **European** | **Middle Eastern** | **South Asian** | **Oceanian** |
| --- | --- | --- | --- | --- | --- | --- | --- | --- |
| American Indian, Native American, Alaska Native | **2** | 12.00  (16.90) | 46.90  (56.90) | 4.90  (3.00) | 25.20  (28.70) | 10.60  (15.00) | 0.44 (0.63) | 0.00  (0.00) |
| Black or African American | **80** | 82.20  (11.90) | 0.56  (1.38) | 0.42 (0.73) | 12.90  (9.23) | 1.98  (3.40) | 1.63 (3.00) | 0.32  (0.48) |
| Hispanic/Latino(a) | **2** | 14.00  (8.79) | 40.70  (20.20) | 0.84  (1.90) | 29.90  (6.21) | 14.50  (6.60) | 0.00  (0.00) | 0.16  (0.22) |
| Middle Eastern or North African/Mediterranean | **1** | 0.46  (NA) | 0.00  (NA) | 0.00  (NA) | 10.60  (NA) | 75.90  (NA) | 13.10 (NA) | 0.00  (NA) |
| More than one category (excluding Hispanic/Latino(a)) | **22** | 39.50  (26.60) | 2.89  (9.18) | 2.43  (9.60) | 48.60  (26.70) | 2.16  (3.80) | 3.92 (13.00) | 0.50  (0.64) |
| More than one category (including Hispanic/Latino(a)) | **4** | 6.58  (7.00) | 24.30  (23.50) | 4.57  (7.10) | 55.50  (26.90) | 6.04  (1.90) | 2.86 (4.20) | 0.17  (0.34) |
| Not Reported | **275** | 35.70  (39.10) | 6.37  (19.90) | 1.16  (7.50) | 52.30  (39.10) | 2.50  (6.10) | 1.74 (5.60) | 0.29  (0.51) |
| Prefer not to answer | **1** | 0.00  (NA) | 0.00  (NA) | 0.00  (NA) | 99.00  (NA) | 0.00  (NA) | 1.00  (NA) | 0.00  (NA) |
| Unknown/none of these fully describe me | **4** | 36.90  (14.00) | 0.19  (0.39) | 1.18  (1.50) | 58.20  (7.80) | 2.37  (4.70) | 0.52 (1.00) | 0.60  (1.11) |
| White or European American | **144** | 0.42  (2.00) | 0.41  (1.23) | 0.50  (3.70) | 94.20  (8.30) | 2.78  (6.50) | 1.46 (1.70) | 0.28  (0.47) |
| Overall | **535** | 32.80  (38.70) | 4.10  (15.50) | 0.96  (6.10) | 57.40  (39.20) | 2.72  (6.70) | 1.75 (5.00) | 0.30  (0.50) |

#### **Supplementary Table 9: Number of positive/diagnosed cases and diagnostic yield (positive cases as % of total) by sex, number of indications, and race/ethnicity of cases stratified by study**

|  | **KidsCanSeq** | **NCGENES 2** | **NYCKidSeq** | **P^3^EGS pediatric** | **P^3^EGS prenatal** | **SouthSeq** |
| --- | --- | --- | --- | --- | --- | --- |
| **Total Number**  **(Diagnostic Yield %)** | 47  (8.2) | 7  (7.3) | 168  (17.5) | 141  (26.7) | 60  (19.0) | 150  (28.0) |
| **Sex** |  |  |  |  |  |  |
| Female | 24  (8.5) | 3  (8.6) | 76  (20.4) | 76  (32.1) | 26  (17.9) | 73  (29.2) |
| Male | 23  (8.0) | 4  (6.6) | 92  (15.6) | 65  (22.3) | 34  (19.9) | 77  (27.0) |
| **Indications** |  |  |  |  |  |  |
| 1 | 34  (7.0) | 2  (6.5) | 92  (14.6) | 73  (25.6) | 24  (15.1) | 41  (22.7) |
| >1 | 13  (15.3) | 5  (7.7) | 76  (22.8) | 68  (28.0) | 36  (22.9) | 109  (30.8) |
| **Race/Ethnicity** |  |  |  |  |  |  |
| American Indian, Native American, Alaska Native | 0  (0.0) | 1  (33.3) | 0  (0.0) | 2  (66.7) | 0  (0.0) | 0  (0.0) |
| Asian | 2  (13.3) | 0  (0.0) | 7  (14.3) | 10  (26.3) | 5  (13.2) | 0  (0.0) |
| Black or African American | 7  (13.2) | 0  (0.0) | 23  (15.0) | 3  (17.7) | 0  (0.0) | 19  (23.8) |
| Hispanic/Latino(a) | 18  (8.4) | 0  (0.0) | 74  (20.0) | 57  (26.0) | 9  (19.6) | 1  (50.0) |
| Middle Eastern or North African/Mediterranean | 0  (0.0) | 0  (0.0) | 1  (16.7) | 4  (18.2) | 1  (33.3) | 0  (0.0) |
| Native Hawaiian/Pacific Islander | 0  (0.0) | 0  (0.0) | 0  (0.0) | 0  (0.0) | 0  (0.0) | 0  (0.0) |
| White or European American | 12  (7.1) | 5  (7.1) | 34  (17.5) | 18  (28.1) | 14  (15.4) | 43  (29.9) |
| Missing | 1  (5.9) | 0  (0.0) | 4  (13.8) | 21  (29.6) | 19  (31.7) | 80  (28.6) |
| More than one category (excluding Hispanic/Latino(a)) | 1  (3.3) | 0  (0.0) | 8  (17.4) | 15  (32.6) | 2  (7.4) | 7  (31.8) |
| More than one category (including Hispanic/Latino(a)) | 6  (9.7) | 1  (25.0) | 17  (15.0) | 11  (25.0) | 10  (20.4) | 0  (0.0) |

#### **Supplementary Table 10: Proportions of Inheritance Patterns Among Positive Cases Across Five CSER Studies.**

The table presents the total number of positive/ diagnosed cases, and the percentages of inheritance patterns categorized as: Autosomal dominant (de novo, inherited, or unknown), Autosomal recessive (compound heterozygous or homozygous), and X-linked (de novo, maternal, or unknown) among positive cases in each study.

|  | **KidsCanSeq** | **NCGENES2** | **NYCKidSeq** | **P^3^EGS** | **SouthSeq** |
| --- | --- | --- | --- | --- | --- |
| **Number of total positive cases in unrelated participants** | **47** | **7** | **168** | **201** | **150** |
| **Mode of Inheritance/ Inheritance pattern** |  |  |  |  |  |
| **Autosomal Dominant (All)** | **93.6**% | **71.4**% | **86.3**% | **68.2**% | **74.3**% |
| Autosomal Dominant (de novo) | 14.9% | 14.3% | 51.8% | 50.2% | 32.7% |
| Autosomal Dominant (Inherited) | 27.7% | 28.5% | 16.7% | 9.0% | 12.7% |
| Autosomal Dominant (Unknown) | 51.5% | 28.5% | 17.9% | 9.0% | 24% |
| **Autosomal Recessive (All)** | **2.1**% | **14.3**% | **4.2**% | **19.9**% | **19.3**% |
| Autosomal Recessive (compound het) | 0% | 14.3% | 0.6% | 10.9% | 10.7% |
| Autosomal Recessive (homozygous) | 2.1% | 0% | 3.6% | 9.0% | 7.3% |
| **X-linked (All)** | **4.3**% | **14.3**% | **8.9**% | **11.9**% | **6.4**% |
| X-linked (de novo) | 2.1% | 14.3% | 4.8% | 7.9% | 0.6% |
| X-linked (Maternal) | 2.1% | 0% | 2.4% | 3.4% | 2.6% |
| X-linked (Unknown) | 0% | 0% | 1.8% | 0.1% | 2.6% |

#### **Supplementary Table 11: Mean of consanguinity coefficients (F) by inheritance pattern in 571 unrelated KidsCanSeq cases**

| **Outcome** | **Inheritance pattern** | **Count** | **Mean F** | **Difference in mean F and (95% confidence interval between positive vs negative+ inconclusive cases by t-test)** |
| --- | --- | --- | --- | --- |
| Positive/ diagnosed | Autosomal dominant de novo | 7 | -0.015 | -0.009 (-0.026 - 0.008) |
|  | Autosomal dominant inherited | 13 | -0.011 | -0.005 (-0.013 - 0.004) |
|  | Autosomal dominant unknown | 24 | -0.004 | 0.002 (-0.003 - 0.007) |
|  | Autosomal recessive (compound heterozygous) | 0 | NA |  |
|  | Autosomal recessive (homozygous) | 1 | 0.057 |  |
|  | X-linked | 1 | -0.011 |  |
| Undiagnosed |  | 525 | -0.006 |  |
| All cases |  | 571 | -0.006 |  |

#### **Supplementary Table 12: Mean of consanguinity coefficients (F) by inheritance pattern in 96 NCGENES 2 cases**

| **Outcome** | **Inheritance pattern** | **N** | **Mean F** | **Difference in mean F and (95% confidence interval between positive vs negative+ inconclusive cases by t-test)** |
| --- | --- | --- | --- | --- |
| Positive/ Diagnosed | Autosomal dominant de novo | 1 | 0.014 | NA |
|  | Autosomal dominant inherited | 2 | 0.032 | 0.008 (-0.044 - 0.061) |
|  | Autosomal dominant unknown | 2 | 0.005 | -0.018 (-0.064 - 0.027) |
|  | Autosomal recessive (compound heterozygous) | 1 | -0.0156 | NA |
|  | Autosomal recessive (homozygous) | 0 | NA | NA |
|  | X-linked | 1 | 0.014 | NA |
| Undiagnosed |  | 89 | 0.024 |  |
| All cases |  | 96 | 0.023 |  |

#### **Supplementary Table 13: Mean of consanguinity coefficients (F) by inheritance pattern in 962 unrelated NYCKidSeq cases**

| **Outcome** | **Inheritance pattern** | **Count** | **Mean F** | **Difference in mean F and (95% confidence interval between positive vs negative+ inconclusive cases by t-test)** |
| --- | --- | --- | --- | --- |
| Positive/ Diagnosed | Autosomal dominant de novo | 87 | -0.006 | -0.003 (-0.007 - 0.014) |
|  | Autosomal dominant inherited | 28 | -0.002 | 0.001 (-0.005 - 0.008) |
|  | Autosomal dominant unknown | 30 | -0.002 | 0.001 (-0.007 - 0.009) |
|  | Autosomal recessive (compound heterozygous) | 1 | -0.003 | NA |
|  | Autosomal recessive (homozygous) | 6 | 0.030 | 0.036 (-0.010 - 0.080) |
|  | X-linked | 15 | -0.002 | 0.005 (-0.002 - 0.012) |
| Undiagnosed |  | 795 | -0.003 |  |
| All cases |  | 962 | -0.003 |  |

#### **Supplementary Table 14: Mean of consanguinity coefficients (F) by inheritance pattern in 844 unrelated P^3^EGS cases**

| **Outcome** | **Inheritance pattern** | **Count** | **Mean F** | **Difference in mean F and (95% confidence interval between positive vs negative+ inconclusive cases by t-test)** |
| --- | --- | --- | --- | --- |
| Positive/ Diagnosed | Autosomal dominant de novo | 101 | -0.006 | -0.01 (-0.015 - -0.004) |
|  | Autosomal dominant inherited | 18 | -0.001 | -0.005 (-0.013 - 0.002) |
|  | Autosomal dominant unknown | 18 | 0.003 | -0.001 (-0.010 - 0.009) |
|  | Autosomal recessive (compound heterozygous) | 22 | -0.010 | -0.014 (-0.030 - 0.001) |
|  | Autosomal recessive (homozygous) | 18 | 0.048 | 0.044 (0.022 - 0.070) ***** |
|  | X-linked | 24 | 0.001 | -0.003 (-0.011 - 0.005) |
| Undiagnosed |  | 643 | 0.004 |  |
| All cases |  | 844 | 0.003 |  |

* Indicates T-statistic *P*-value < 0.002.

#### **Supplementary Table 15: Mean of consanguinity coefficients (F) by inheritance pattern in 537 SouthSeq cases**

| **Outcome** | **Inheritance pattern** | **N** | **Mean F** | **Difference in mean F and (95% confidence interval between positive vs negative+ inconclusive cases by t-test)** |
| --- | --- | --- | --- | --- |
| Positive/ Diagnosed | Autosomal dominant de novo | 43 | -0.003 | 0.002 (-0.005 - 0.008) |
|  | Autosomal dominant inherited | 18 | -0.007 | -0.002 (-0.008 - 0.003) |
|  | Autosomal dominant unknown | 30 | -0.006 | -0.002 (-0.010 - 0.006) |
|  | Autosomal recessive (compound heterozygous) | 16 | -0.003 | 0.001 (-0.008 - 0.010) |
|  | Autosomal recessive (homozygous) | 11 | 0.000 | 0.004 (-0.007 - 0.016) |
|  | X-linked | 7 | 0.000 | 0.004 (-0.014 - 0.022) |
| Undiagnosed |  | 387 | -0.005 |  |
| All cases |  | 537 | -0.005 |  |

#### **Supplementary Table 16: P/LP variant type frequencies in 4 CSER studies and chi-square test of 4 sites with 4 different variant types (df=9)**

| **P/LP Variant type** | **All** | **KidsCanSeq** | **NYCKidSeq** | **P^3^EGS** | **SouthSeq** | **Chi-Square (*P*-value)** |
| --- | --- | --- | --- | --- | --- | --- |
| frameshift | **129**  **(22.6%)** | 17  (36.2%) | 31  (21.4%) | 53  (24.7%) | 28  (17.1%) | 19.15  (0.024) * |
| splice site | **34**  **(6%)** | 6  (12.8%) | 3  (2.1%) | 10  (0.5%) | 15  (9.1%) |  |
| missense | **196**  **(34.3%)** | 12  (25.5%) | 45  (31.0%) | 92  (42.8%) | 47  (28.7%) |  |
| stop-gain | **110**  **(19.3%)** | 12  (25.5%) | 19  (13.1%) | 54  (25.1%) | 25  (15.2%) |  |
| **Sub-total** | **469**  **(82.1%)** | **47**  **(100%)** | **98**  **(67.6%)** | **209**  **(97.2%)** | **115**  **(70.1%)** |  |
| start lost | **1**  **(0.2%)** | 0  (0%) | 0  (0%) | 1  (0.5%) | 0  (0%) |  |
| in-frame deletion | **5**  **(0.9%)** | 0  (0%) | 3  (2.1%) | 2  (0.9%) | 0  (0%) |  |
| in-frame insertion | **1**  **(0.2%)** | 0  (0%) | 0  (0%) | 0  (0%) | 1  (0.6%) |  |
| CNV | **90**  **(15.8%)** | 0  (0%) | 40  (27.6%) | 2  (0.9%) | 48  (29.3%) |  |
| insert-delete | **5**  **(0.9%)** | 0  (0%) | 4  (2.8%) | 1  (0.5%) | 0  (0%) |  |
| **All-total** | **571**  **(100%)** | **47**  **(100%)** | **145**  **(100%)** | **215**  **(100%)** | **164**  **(100%)** |  |

* Chi-Square statistic and *P*-value of the 4x4 table (9 df) of the 4 centers and 4 variant types: frameshift, splice site, missense, stop-gain.

There were 4 P/LP variants in NCGENES 2, 1 each of start lost, frameshift, missense, stop gain.

#### **Supplementary Table 17: Variants with maximum population specific allele frequency =>0.001 in gnomAD V4**

| **dbSNP Variant**  **ID** | **Allele Frequency of variants in gnomAD V4 Populations** | | | | | | | | **Inheritance Pattern** |
| --- | --- | --- | --- | --- | --- | --- | --- | --- | --- |
|  | Admixed American | Finnish | Middle Eastern | European  (Non-Finnish) | South Asian | Ashkenazi Jewish | East Asian | African/ A.A* |  |
| [rs1801155](http://www.ncbi.nlm.nih.gov/snp/rs1801155) | 0.00058 | 0 | 0.022 | 0.00041 | 0.00053 | 0.037 | 0 | 0.00012 | Autosomal dominant (Inherited) |
| [rs61749370](https://www.ncbi.nlm.nih.gov/snp/rs61749370) | 0.001 | 0.0045 | 0.00033 | 0.0045 | 0.0004 | 0.0033 | 0 | 0.0002 | Autosomal dominant (Unknown) |
| [rs80338794](http://www.ncbi.nlm.nih.gov/snp/rs80338794) | 0.000017 | 0.0046 | 0 | 0.00023 | 0.000011 | 0 | 0 | 0.000013 | Autosomal recessive (Heterozygous) |
| [rs76723693](http://www.ncbi.nlm.nih.gov/snp/rs76723693) | 0.0012 | 0 | 0 | 1.117E-06 | 0 | 0 | 0 | 0.0037 | X-linked  (Maternal) |
| [rs146013446](http://www.ncbi.nlm.nih.gov/snp/rs146013446) | 0.00012 | 0.00082 | 0.00033 | 0.0013 | 0.00018 | 0 | 0 | 0.00026 | Autosomal recessive (Heterozygous) |
| [rs138659167](http://www.ncbi.nlm.nih.gov/snp/rs138659167) | 0.002 | 0.0016 | 0.00033 | 0.0092 | 0.00013 | 0.012 | 0 | 0.0015 | Autosomal recessive (Heterozygous) |
| [rs121965020](http://www.ncbi.nlm.nih.gov/snp/rs121965020) | 0.00017 | 0.0021 | 0 | 0.00053 | 0 | 0 | 0 | 0.000067 | Autosomal recessive (Homozygous) |
| [rs80338939](http://www.ncbi.nlm.nih.gov/snp/rs80338939) | 0.0052 | 0.0095 | 0.0041 | 0.0083 | 0.0006 | 0.0036 | 0.000022 | 0.0013 | Autosomal recessive (Heterozygous) |

*A.A – African American

#### **Supplementary Table 18:** **Recurrent diagnostic pathogenic variants found among 3008 unrelated cases across 5 CSER studies**

| **Gene (Number of recurrent variants)** | **Nucleotide change and position** | **Inheritance pattern** | **CSER center** | **Human Genome reference version** |
| --- | --- | --- | --- | --- |
| *CDK13* (2) | c.2525A>G  (p.Asn842Ser) | Autosomal dominant (De novo) | P^3^EGS | Hg37 |
| *FGFR3* (2) | c.742C>T  (p.Arg248Cys) | Autosomal dominant (De novo), Autosomal dominant unknown | P^3^EGS | Hg37 |
| *PTPN11* (2) | c.317A>C  (p.Asp106Ala) | Autosomal dominant (Maternal), Autosomal dominant (Paternal) | SouthSeq,  NYCKidSeq | Hg38 |
| *PTPN11* (2) | c.182A>G  (p.Asp61Gly) | Autosomal dominant (De novo) | P^3^EGS | Hg37 |
| *PTPN11* (2) | c.854T>C  (p.Phe285Ser) | Autosomal dominant (De novo) | P^3^EGS | Hg37 |
